# Patterns of age disparities in colon and lung cancer survival: a systematic narrative literature review

**DOI:** 10.1101/2020.09.08.20190231

**Authors:** Sophie Pilleron, Helen Gower, Maryska Janssen-Heijnen, Virginia Signal, Jason Gurney, Eva JA Morris, Ruth Cunningham, Diana Sarfati

**Author notes:** **Corresponding author:** Sophie Pilleron, Department of Public Health, University of Otago, PO Box 7343, Wellington, New Zealand.; /.

## Abstract

**Objective:** To identify patterns of age disparities in cancer survival, using colon and lung cancer as exemplars.

**Methods:** We conducted a systematic review of literature published in EMBASE, MEDLINE, Scopus, and Web of Science according to PRISMA guidelines. We included population-based studies in patients with colon or lung cancer. We assessed the quality of included studies against selected evaluation domains from the QUIPS Tool, and items concerning statistical reporting. We evaluated age disparities using the absolute difference in survival or mortality rates between middle-aged group and the oldest age group, or by describing survival curves.

**Results:** Out of 2,162 references reviewed, we retained 35 studies (15 for colon, 18 for lung, 2 for both sites). Regardless of the cancer site, included studies were highly heterogeneous and often of poor quality. The magnitude of age disparities in survival varied greatly by sex, ethnicity, socio-economic status, stage at diagnosis, cancer site and morphology, the number of nodes examined, and by treatment strategy. Although results were inconsistent for most characteristics, we consistently observed greater age disparities for females with lung cancer compared to males. Also, age disparities increased with more advanced stages for colon cancer, and decreased with more advanced stages for lung cancer.

**Conclusions:** Although age is one of the most important prognostic factors in cancer survival, age disparities in colon and lung cancer survival have so far been understudied in population-based research. Further studies are needed to better understand age disparities in colon and lung cancer survival. (PROSPERO registration number: CRD42020151402).

**Article Summary:** *Strengths and limitations of this study:*

- For the first time, we conducted a systematic review of population-based studies relating to differences in cancer survival between middle-aged and older patients, using colon and lung cancer as exemplar cancers.
- We limited our search to peer-reviewed original articles and letters to Editors published in English up until 30 September 2019.
- We excluded clinical studies and trials because of the strict selection of patients and the common underrepresentation of older patients in these studies.
- We could not conduct any quantitative analysis (such as meta-analysis) because of the vast heterogeneity of studies included, which prevented us from quantifying the relationship between increasing age and cancer survival.

## Introduction

Poorer cancer survival among older patients has been well documented^1–6^. Although patients with cancer are increasingly surviving their disease thanks to advances in treatment^2–6^, those who are older have not benefitted from these advances to the same magnitude as their middle-aged counterparts, widening the age-related cancer survival gap^2,5,7^.

From a clinical point of view, cancer management in older patients may be different to that of middle-aged patients due to higher comorbidity levels, polypharmacy, age-related physiological changes, and reduced life expectancy^8^. In addition, older adults with cancer are often excluded from randomised clinical trials, limiting the evidence they provide in relation to the benefits and risks of different treatment strategies at older ages^9,10^. Cancer management may also be hindered in older cancer patients by social factors such as reduced social support^11,12^ or healthcare system-related factors such as access to care facilities.

A recent systematic review found that advanced age, low income, low socioeconomic status, presence of comorbidities, advanced stage, and poor tumor grade were associated with lower survival among older adults with cancer, while female gender and being married were associated with better survival^13^. However, the authors did not explore inequalities in cancer survival between age groups, and they excluded studies that included middle-aged patients. They also did not focus on any particular cancer sites. This is important, as it is likely that many factors at different levels (e.g. patient level, health care system level) influence age disparities in cancer survival, and they may vary depending on cancer site.

Worldwide, colon and lung cancers are the most common cancer types diagnosed among adults aged 65 years and older^14^. These two cancer sites have different biology, risk factors and survival outcomes, with colon cancer having a higher five-year relative survival than lung cancer, ranging from 59%-71% for colon cancer and 15%- 22% for lung cancer in high-income countries^7^. These cancers also have a different pattern of age inequalities in survival over time. In colon cancer, disparities in cancer survival between older and younger adults is mainly observed in the first year following diagnosis, while in lung cancer, the excess mortality in older adults is mainly observed after five years of follow-up^5,15^.

To our knowledge, there has been no attempt to summarise the available literature on age disparities in cancer survival. Thus, in this manuscript we conducted a systematic review of studies that have investigated differences in cancer survival between middle-aged and older patients, using the diverse contexts of colon and lung cancer as exemplars. We aimed to identify i) patterns of age-related disparities based on patient and clinical characteristics and ii) the potential gaps in knowledge to inform future research.

## Methods and materials

We conducted a systematic literature search of EMBASE, MEDLINE, Scopus, and Web of Science. Using a Boolean approach, we searched for articles including the following keywords: cancer, colon, lung, survival, and older patients. **Supplementary Table 1** shows the search terms that were used. The search strategy was first set up in EMBASE **(Supplementary Table 2)**, and then adapted for the other databases.

We retained all original articles or letters published in English up until 30 September 2019 that included patients diagnosed with colon or lung cancer. Eligible studies were required to report survival across several age groups (of which at least one was over the age of 65), or investigate the impact of increasing age on survival stratified by at least one other characteristic (e.g. sex, treatment, etc.). We included population-based studies only. We excluded clinical studies and trials because of their strict inclusion criteria and the underrepresentation of older adults^9^. The PICO criteria for our review are shown in **Supplementary Table 3**.

### Study selection

We selected eligible articles using a three-step process: (i) after removal of duplicate records, SP screened all titles to remove irrelevant studies, with a 10% random sample of these verified by VS. (ii) For each study retained after title screening, SP screened all abstracts, with a 10% random sample of these checked by HG. (iii) The full text of all retained papers were retrieved and assessed twice for eligibility by SP, with a 10% random sample verified by HG. **Supplementary Table 4** lists all references not included in the final selection after screening the full text, along with the justification of their exclusion. In addition, SP scanned the reference lists of all included studies for additional relevant studies. If one of the authors knew a study that met the eligibility criteria, we included it if relevant. The origin of the studies (i.e. database search or reference lists) are specified in **Supplementary Table 4** for excluded papers, and in **Table 1** for included papers.

**Table 1.**
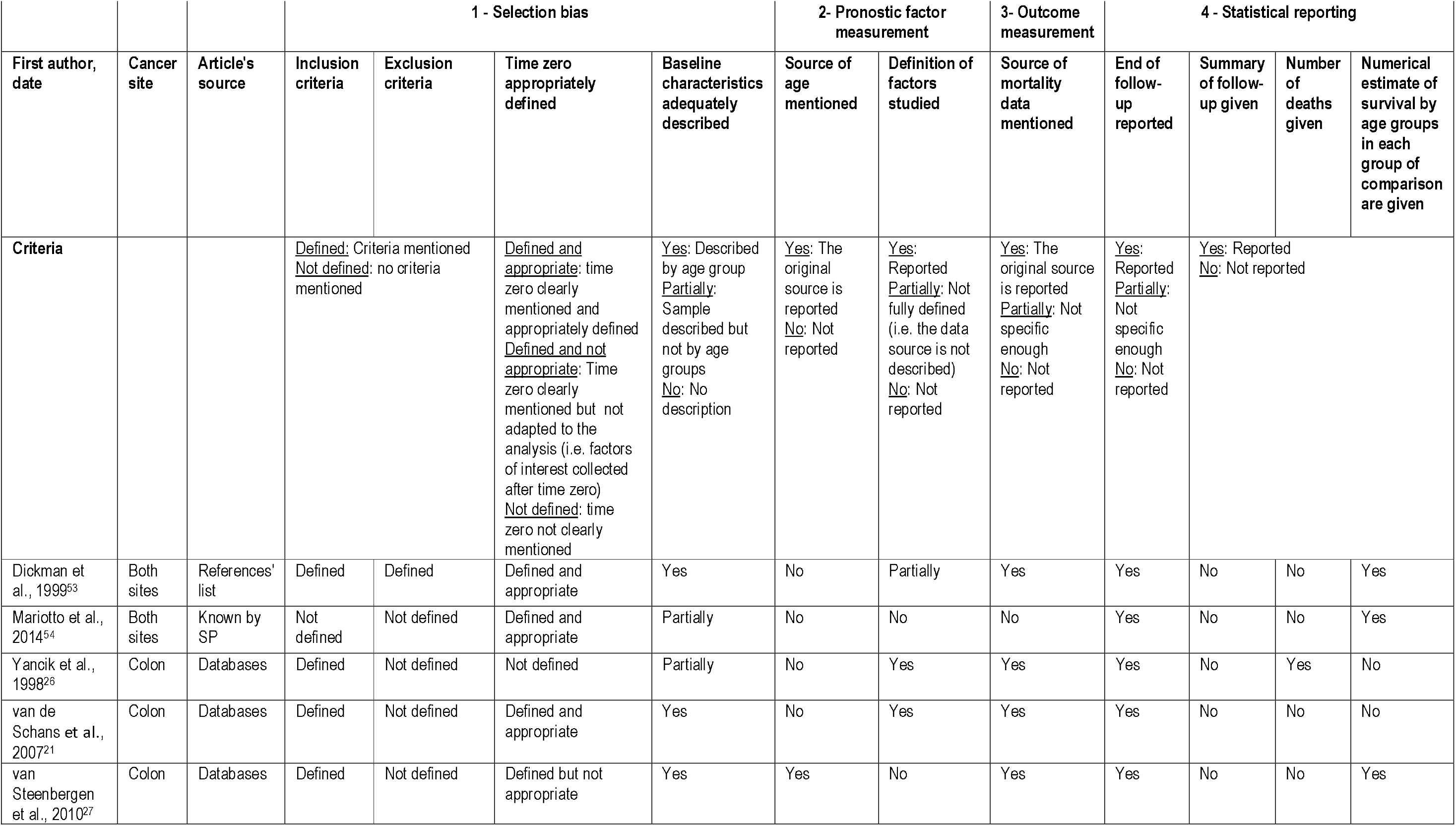
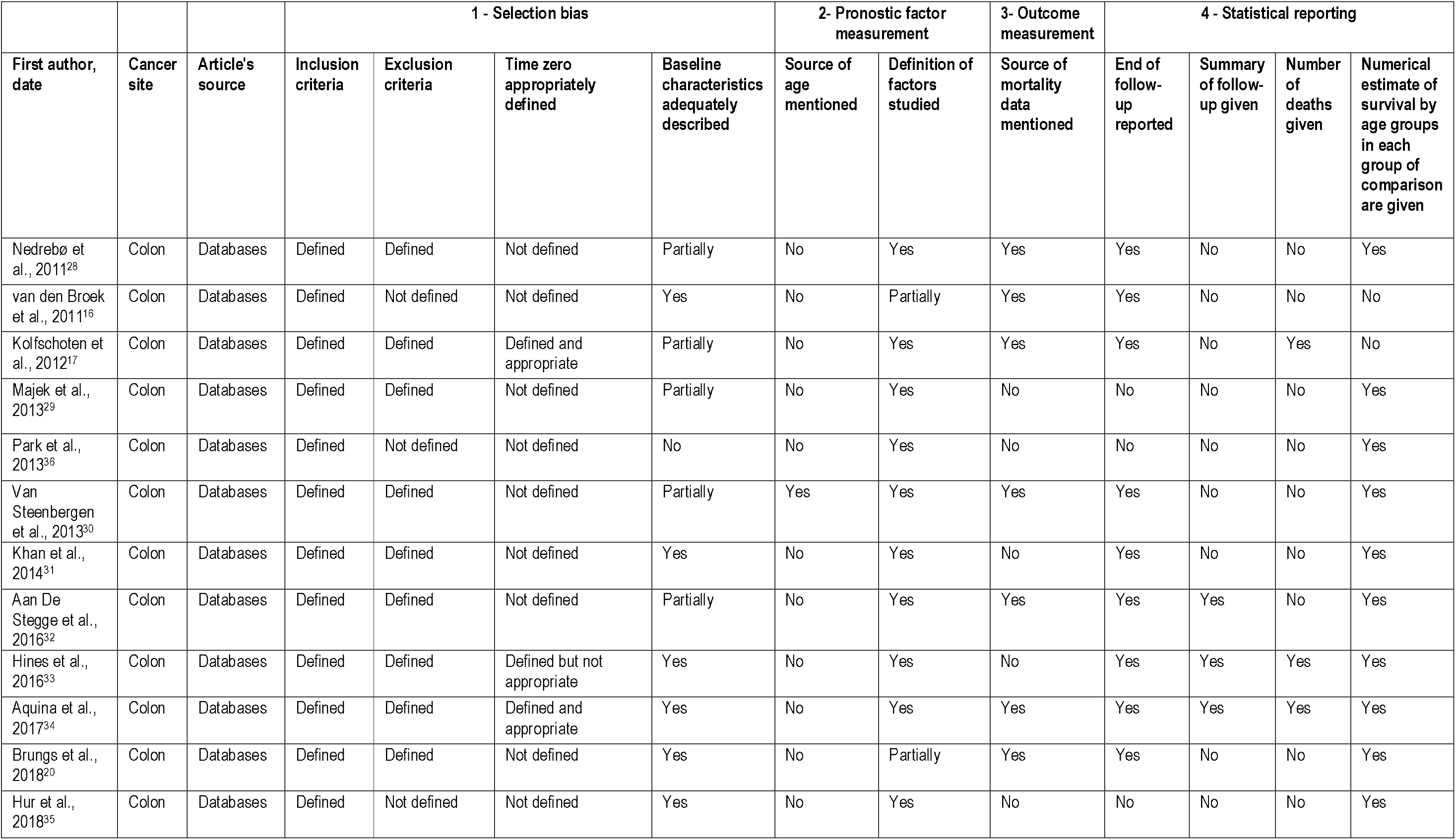
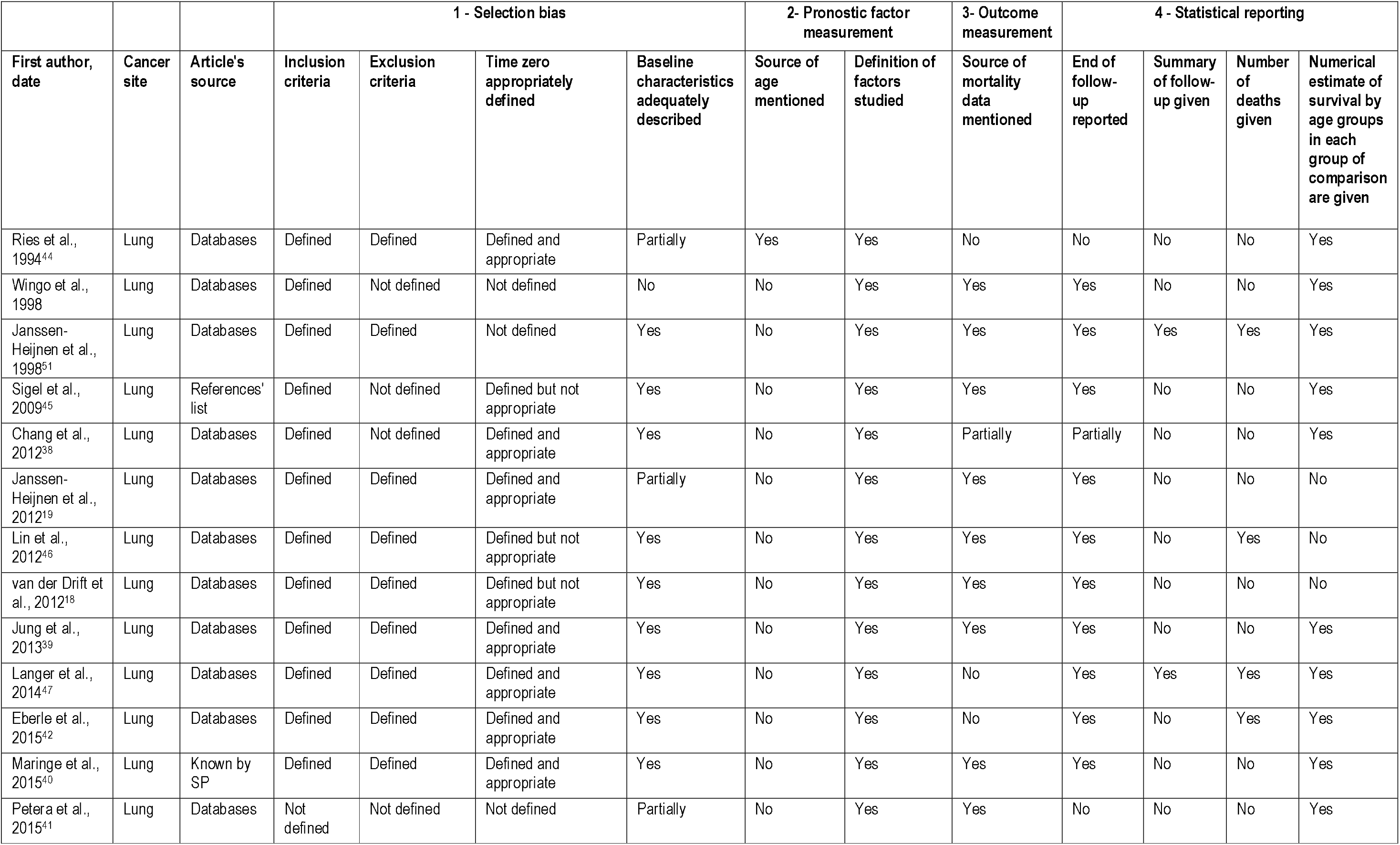
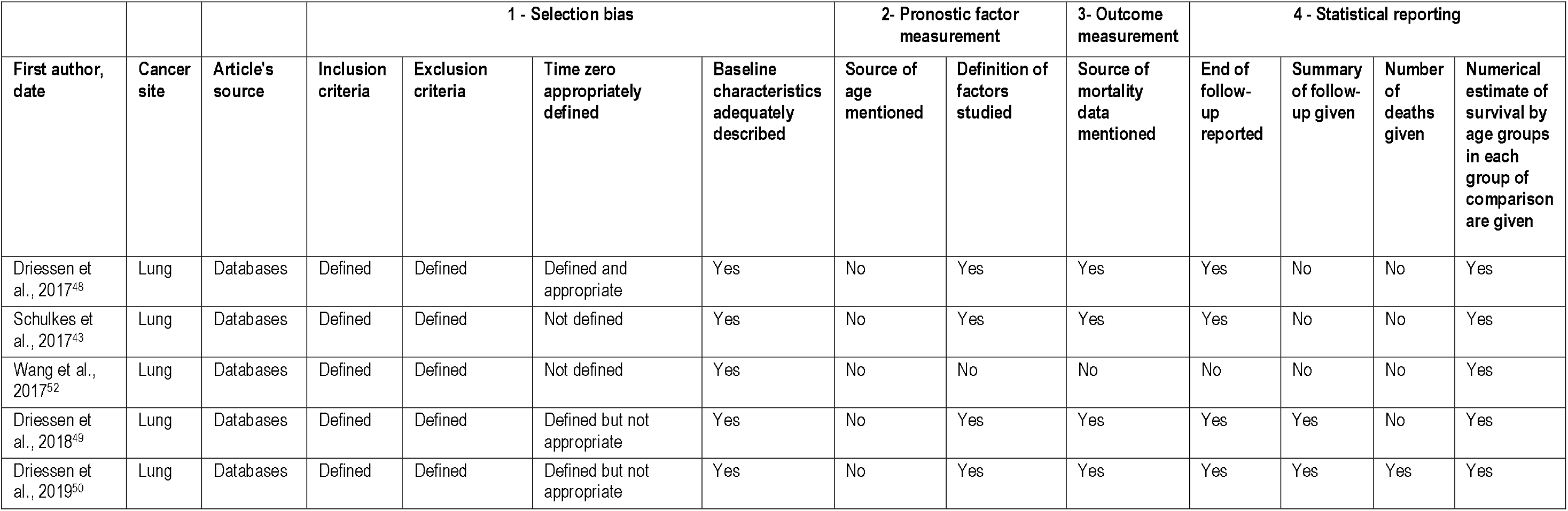
Quality assessment of included studies

### Data collection process and data items

For all included studies, SP and HG independently extracted the following information: first author; year of publication; location of data; study objective; cancer type; stage at diagnosis; age at diagnosis; exclusion criteria; cancer diagnosis period; source of cancer data; source of mortality data; measure of age; source of age; sampling; time origin; end of follow-up; survival/mortality metrics; method; sample size; time of follow-up; number of deaths; characteristic(s) studied and their definition.

In cases where an eligible study contained no numerical survival estimates but presented one or more graphs showing survival by age group stratified by another characteristics (e.g. sex, stage at diagnosis, etc.), SP emailed the corresponding author to request numerical data^16–21^.

SP and HG independently assessed the quality of included studies against selected evaluation domains from the QUIPS Tool^22^: study participation; prognostic factor measurement; outcome measurement; and statistical reporting. We adapted the items within each domain to our study. Also, we used selected items among those suggested by Altman et al. to assess statistical reporting^23^.

Where numerical survival estimates were available, we assessed age disparities in survival by calculating the absolute difference in (overall or relative) survival between middle-aged patients (age groups differed depending on the availability of data) and the oldest age group, to give a sense of trends and inform discussion. When survival estimates were available for several periods of cancer diagnosis, we retained estimates for the latest period. Where numerical survival estimates were not available, we described survival curves by age group and the characteristic(s) of interest. For mortality rates, we computed the absolute difference between the mortality rate in the oldest age group with that in the middle-aged age group, again to give a sense of trends and inform discussion. We reported confidence intervals or p-values when available.

We collected and logged references in Zotero 5.0.73. We used the Rayyan free web application for the title and abstract screening^24^. The Preferred Reporting Items for Systematic Reviews and Meta-Analyses (PRISMA) guidelines were used for the review^25^, and we registered our review protocol in the International Prospective Register of Systematic Review (PROSPERO registration number: CRD42020151402).

### Patient and public involvement

No patient involved.

## Results

We screened 2,162 references for eligibility and retained 35 studies **(Figure 1)**: fifteen studies on colon cancer survival^16,17,20,21,26–36^, eighteen on lung cancer survival^18,19,37–52^ and two studies on both colon and lung cancer survival^53,54^.

### Quality assessment (Table 1)

Essential information to appropriately interpret survival analysis results (*i.e*. the number of events, end of followup, numerical estimates of survival) were missing in a substantial proportion of the included studies. For example, fifteen studies did not report the time origin from which the survival time had been calculated^16,20,26,28–32,35–37,41,43,51,52^, and six studies did not indicate the end of follow-up date^29,35,36,41,44,52^. In 28 articles the authors did not report follow-up time^16–21,26–31,35,37–44,46,48,52–54^, and the number of deaths were missing in 26 artides^16,18–21,27–32,35,37–41,43,44,48,49,52–54^ only three studies reported the source of age at diagnosis (from medical records)^27,30–44^. In seven studies, the authors did not provide numerical survival estimates^16–19,21,26,46^.

### Characteristics of included studies (Tables 2–5)

All studies used population-based cancer registry data. Only one study analysed a random sample of patients^26^. Of seventeen studies examining colon cancer, five studies aimed at investigating age disparities in colon cancer survival **(Table 2)**^16,17,31,32,34^. Six studies used data from The Netherlands^16,17,21,27,30,32^, and five presented data from the U.S.A.^26,31,33,34–54^. The remaining studies used data from Finland^53^, Norway^28^, Germany^29^, Korea^35,36^, and Australia^20^. Ten studies included all cancer stages^16,17,21,26,28,29,35,36,53,54^, three studies restricted their analyses to stage III cancer^20,27,33^, and four to stages l-lll^30–32,34^. Nine studies included all patients whatever their age at diagnosis^16,17,27,28,32,34–36,53^, with the inclusion criterion for age varying widely in the remaining studies. All studies analysed the age at diagnosis using age categories but the number and boundaries of these varied across studies **(Table 4)**. Eight studies presented relative survival (RS) estimates^16,28–30,35,36,53,54^ and five studies presented overall survival (OS) estimates **(Table 4)**^20,21,26,27,32^. The remaining studies showed 30-day postoperative mortality rates^17^, the cumulative incidence of death at five years^31^, or mortality rates^33,34^.

**Table 2.**
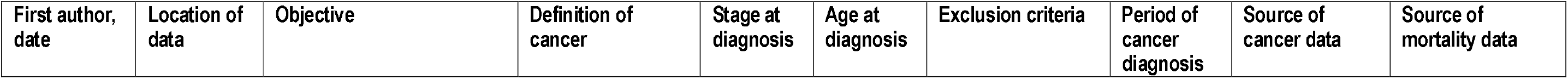
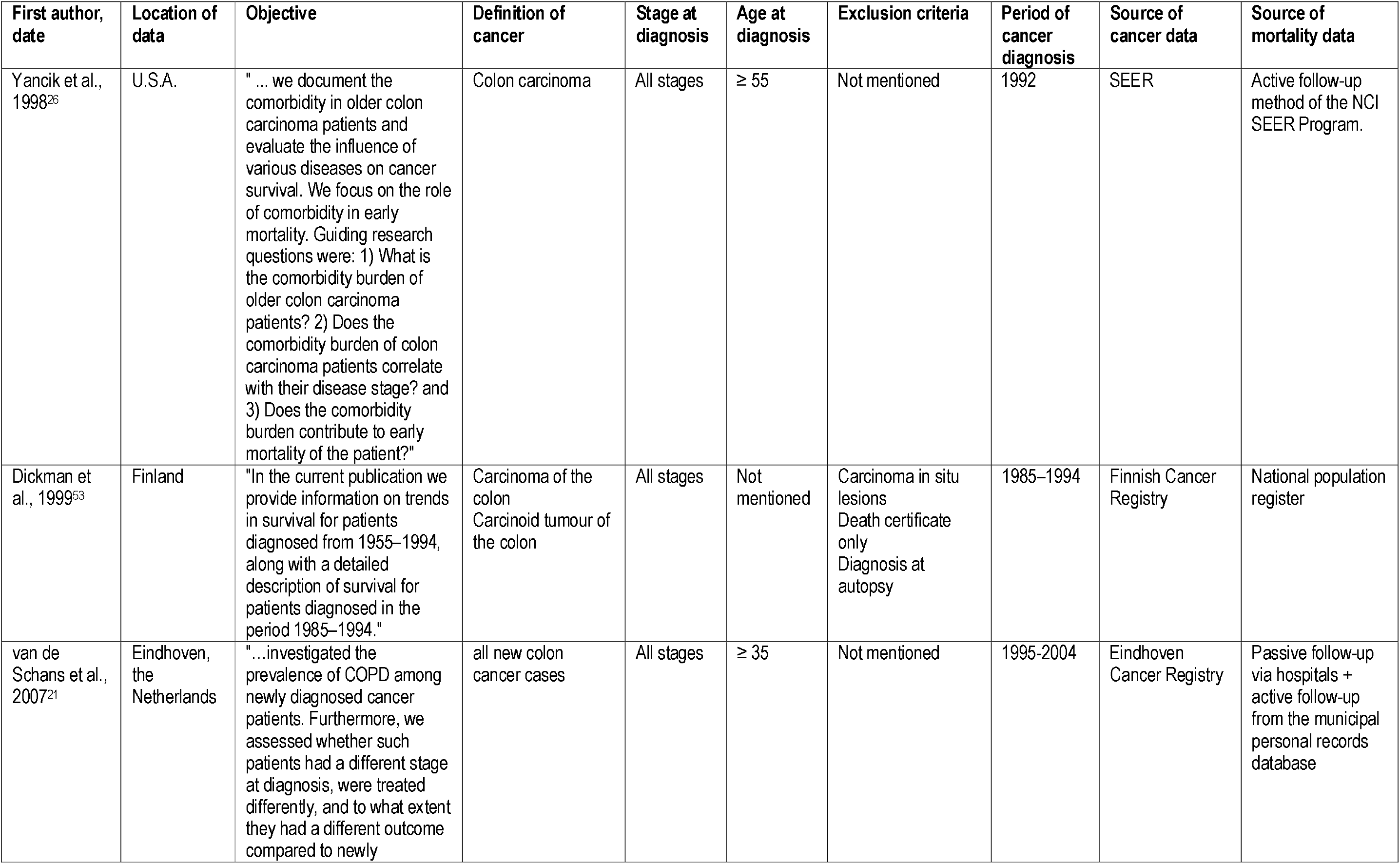
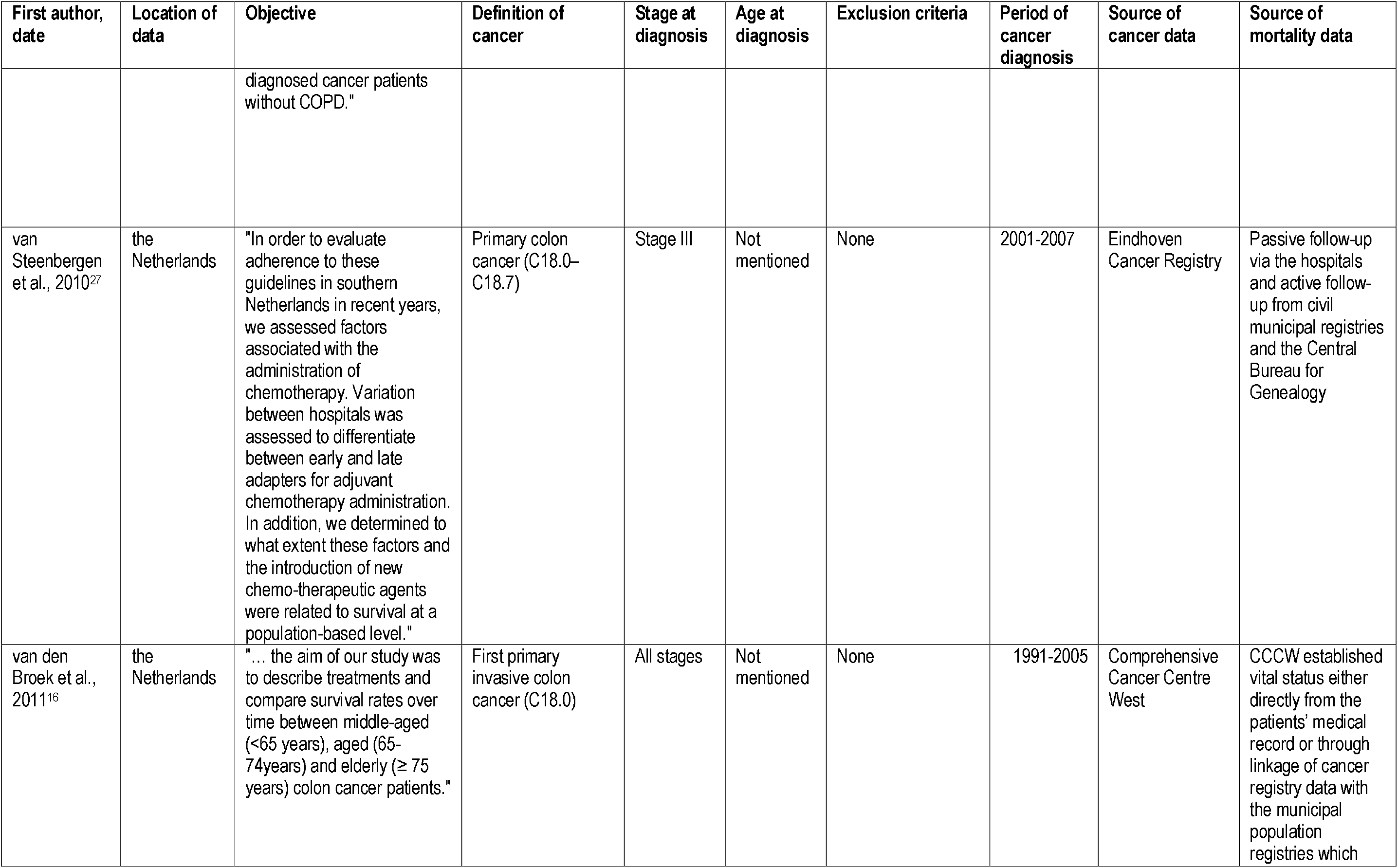
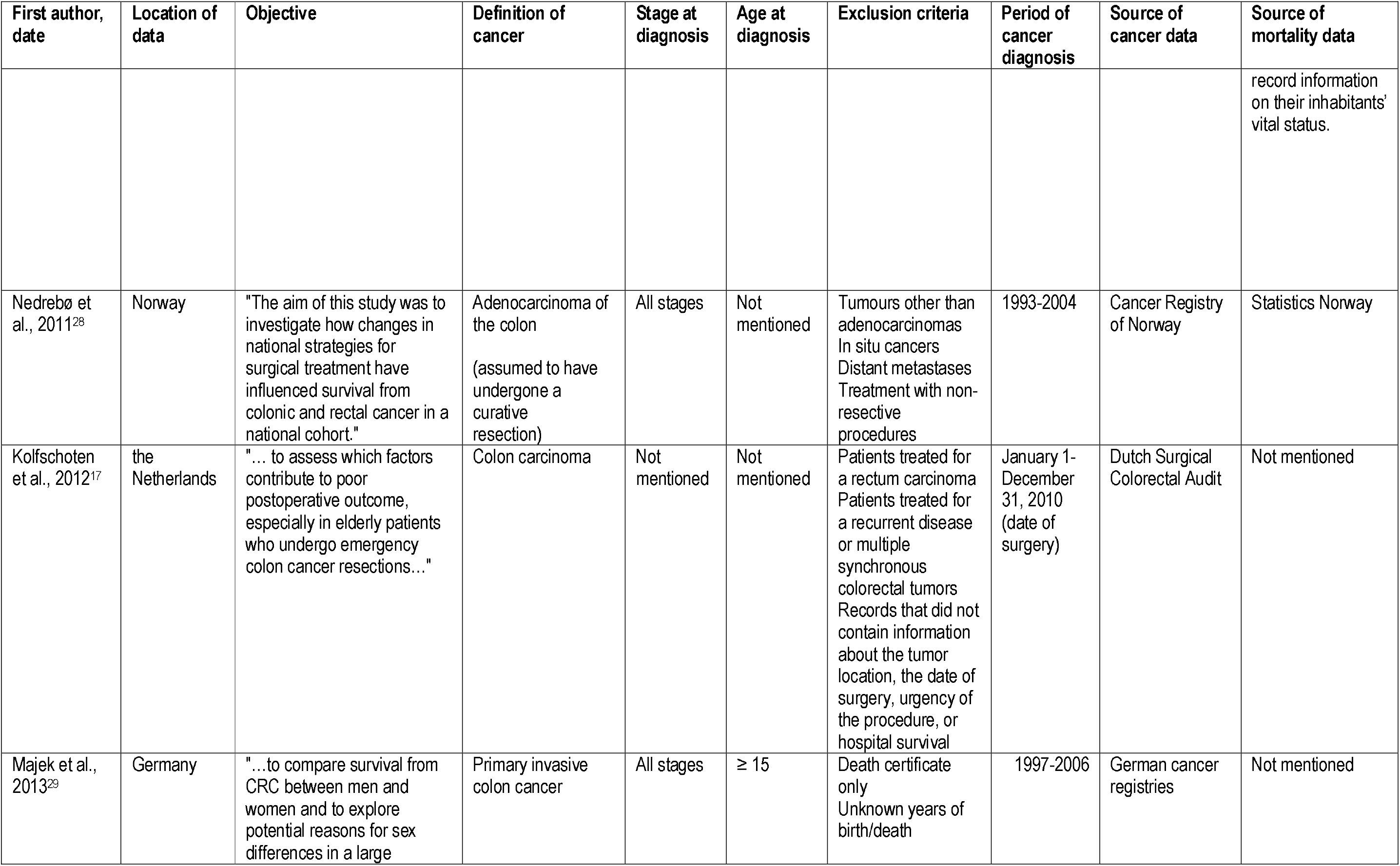
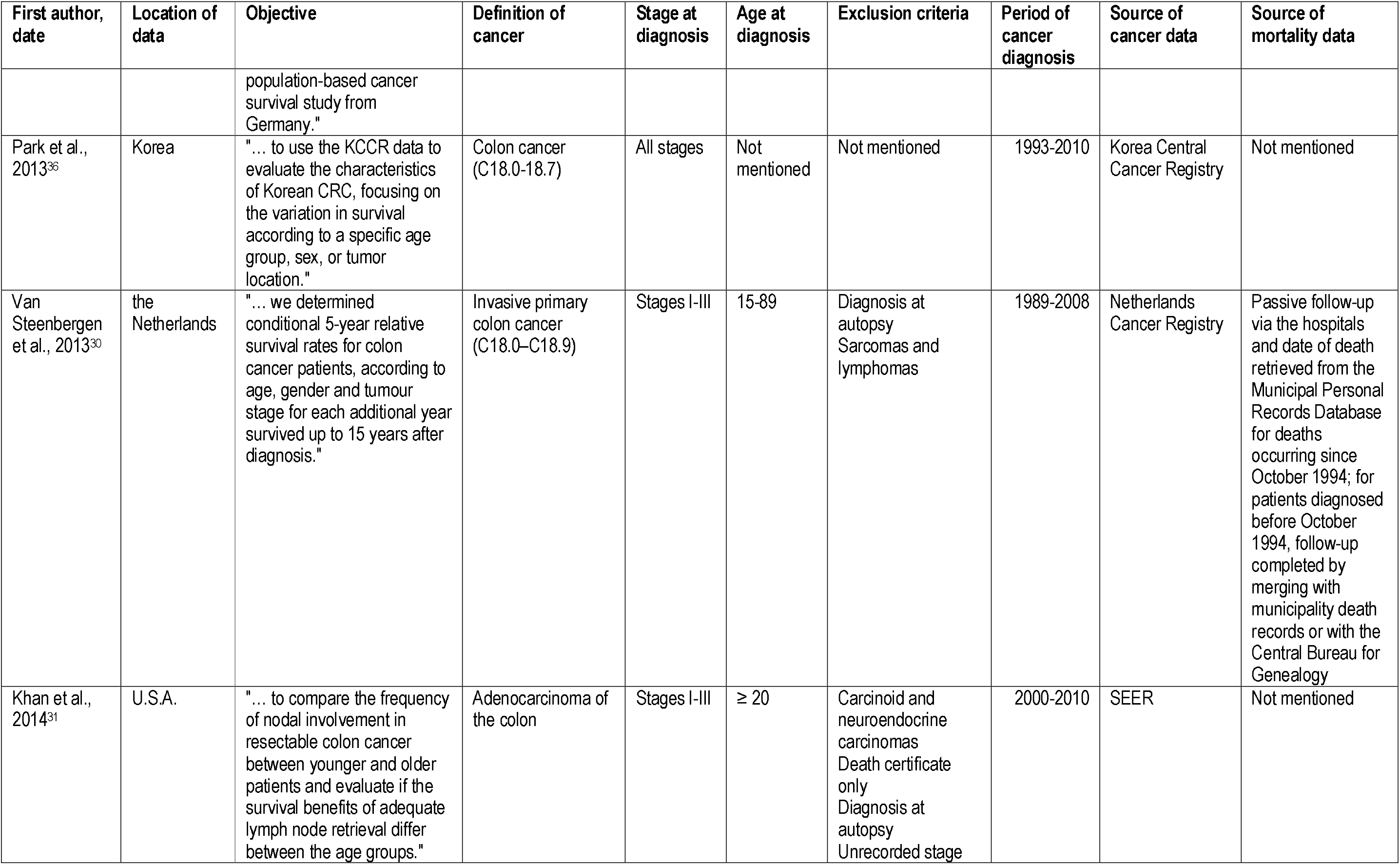
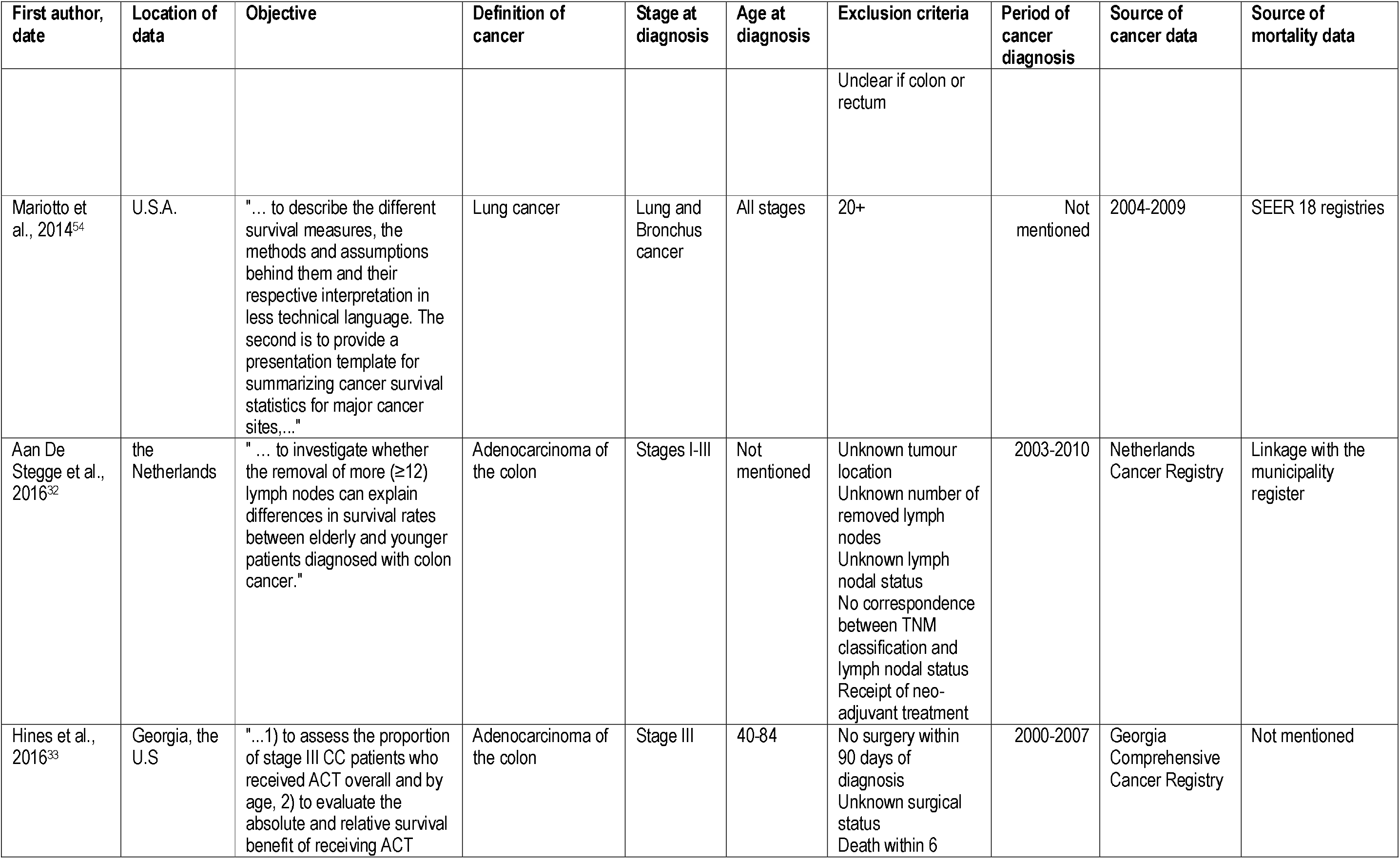
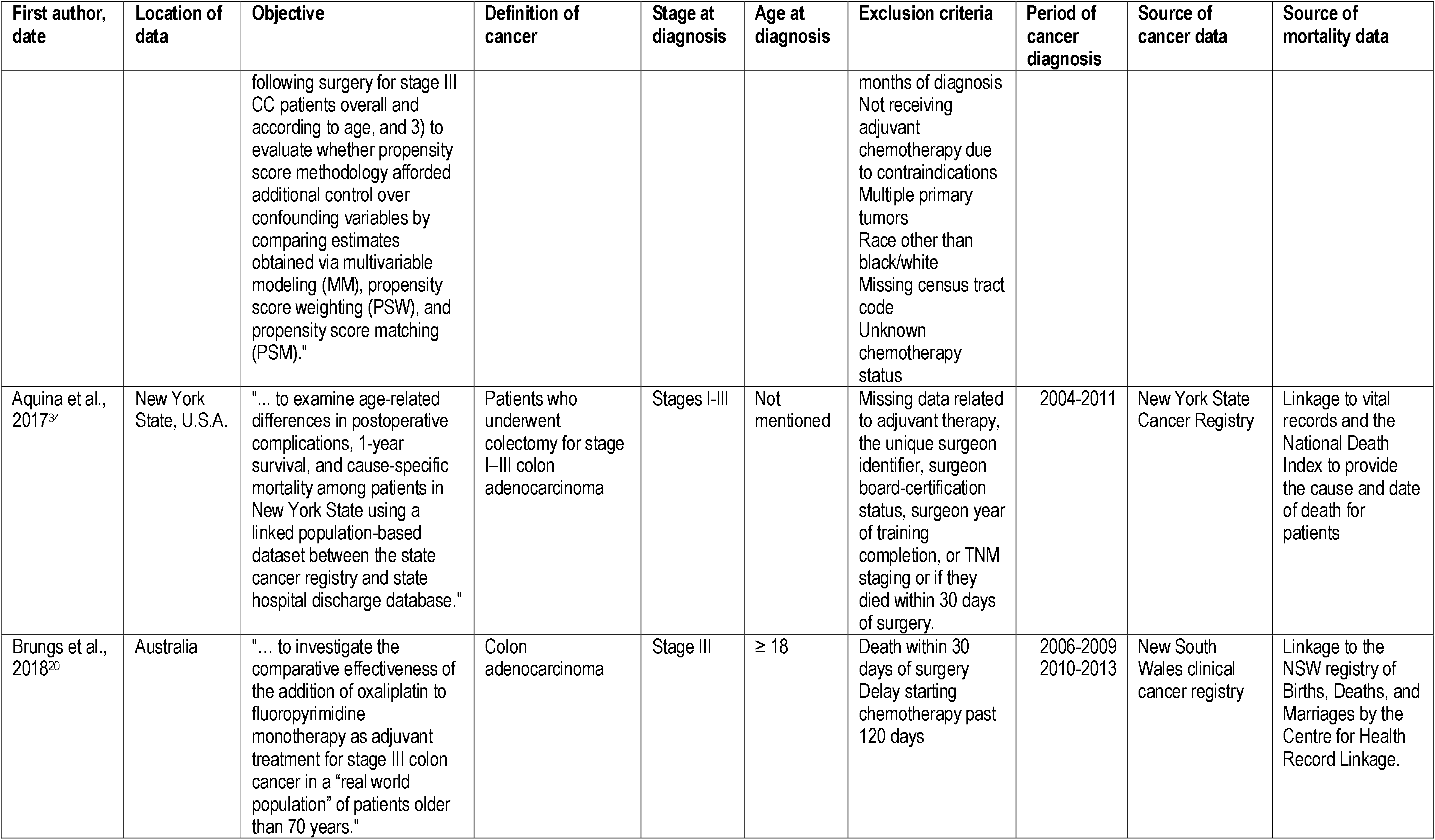
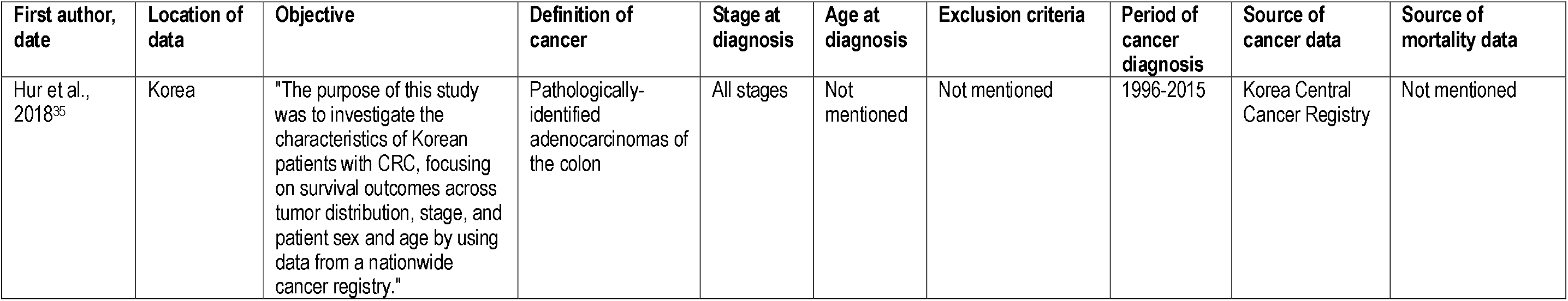
Characteristics of included studies about colon cancers

**Table 4.**
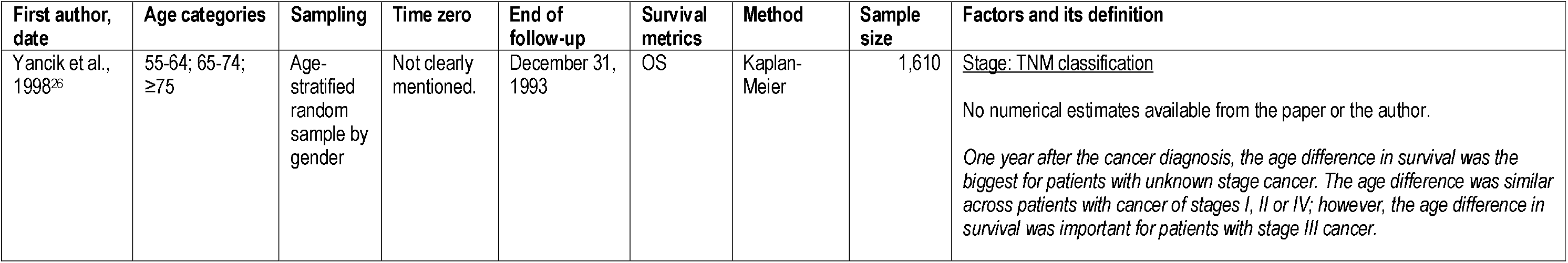
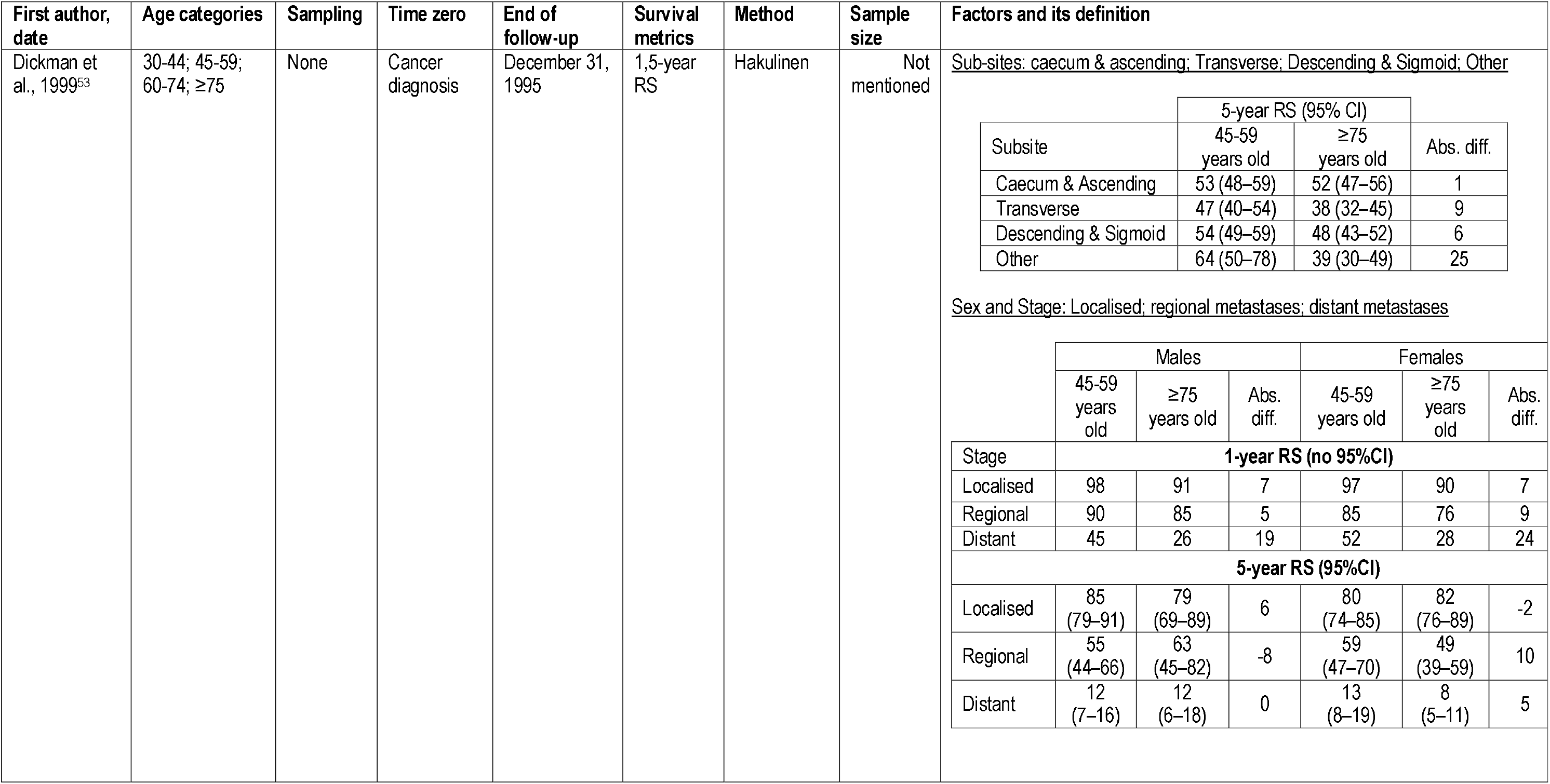
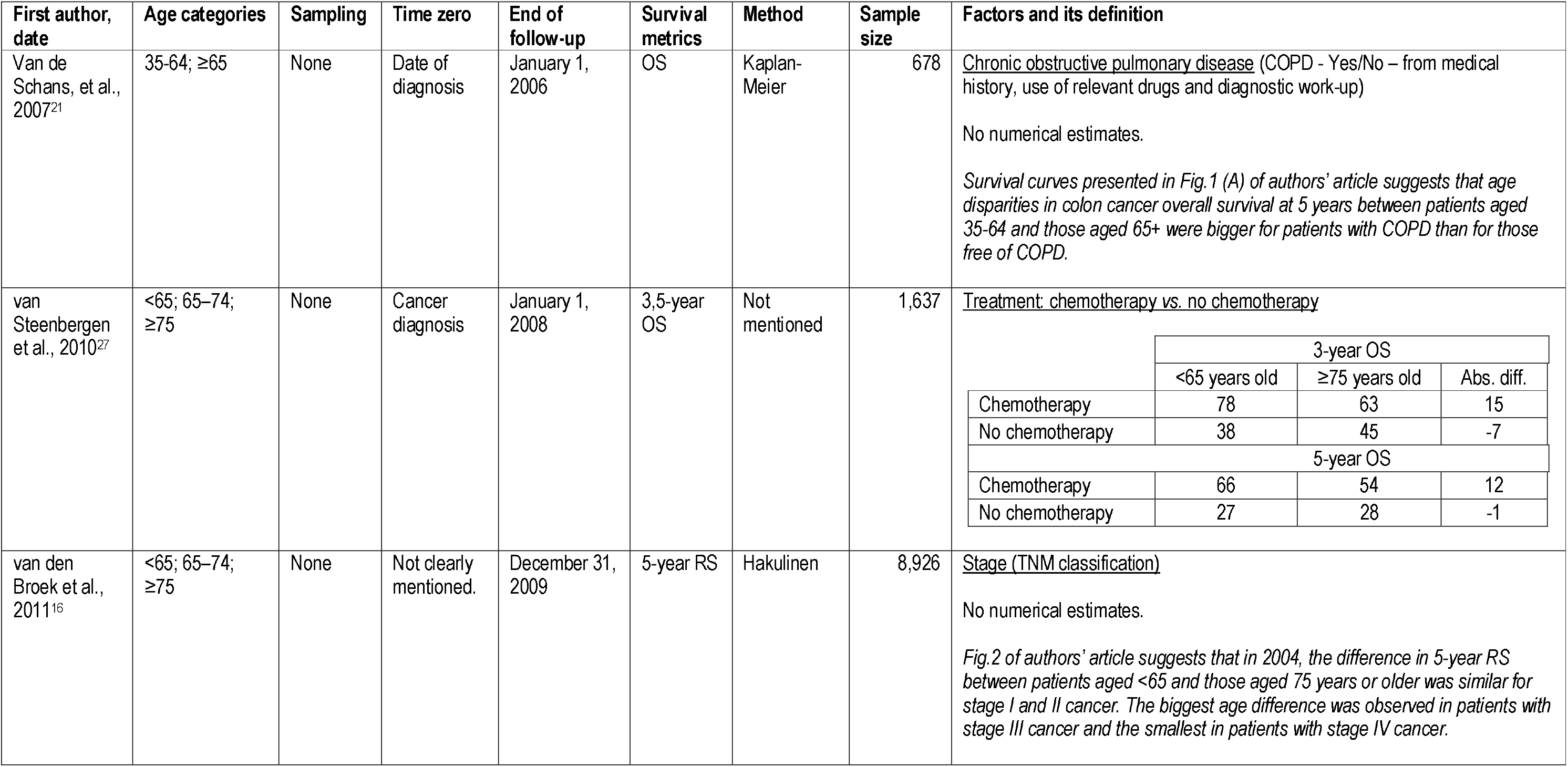
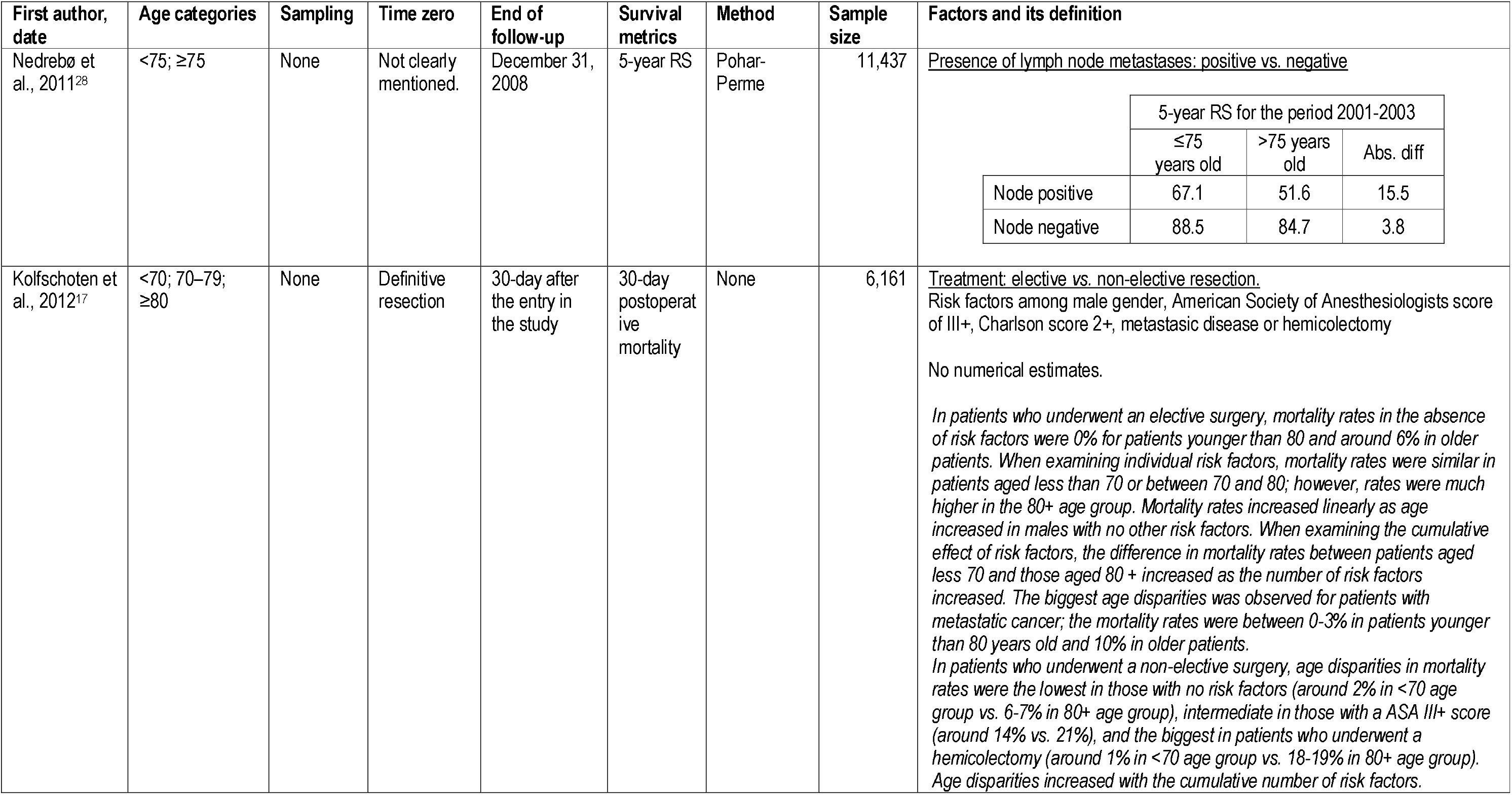
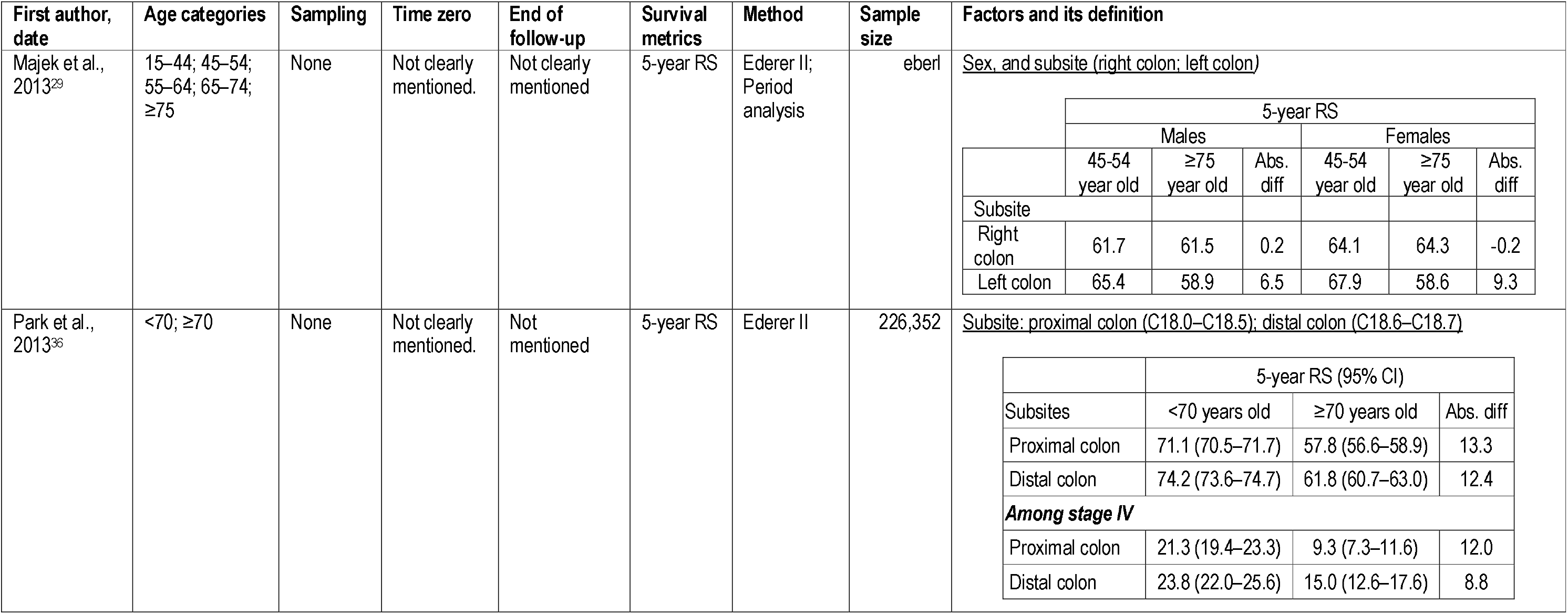
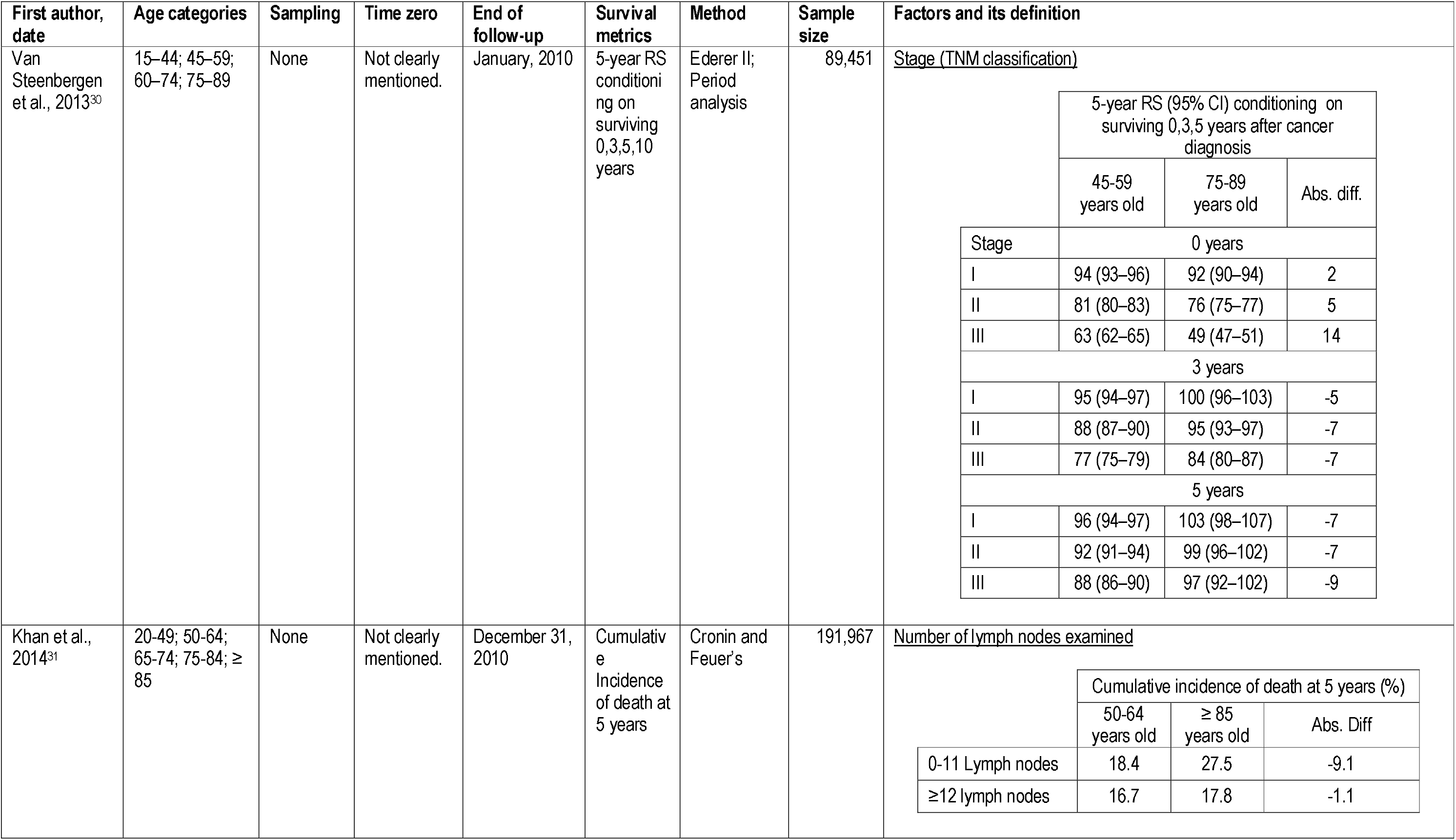
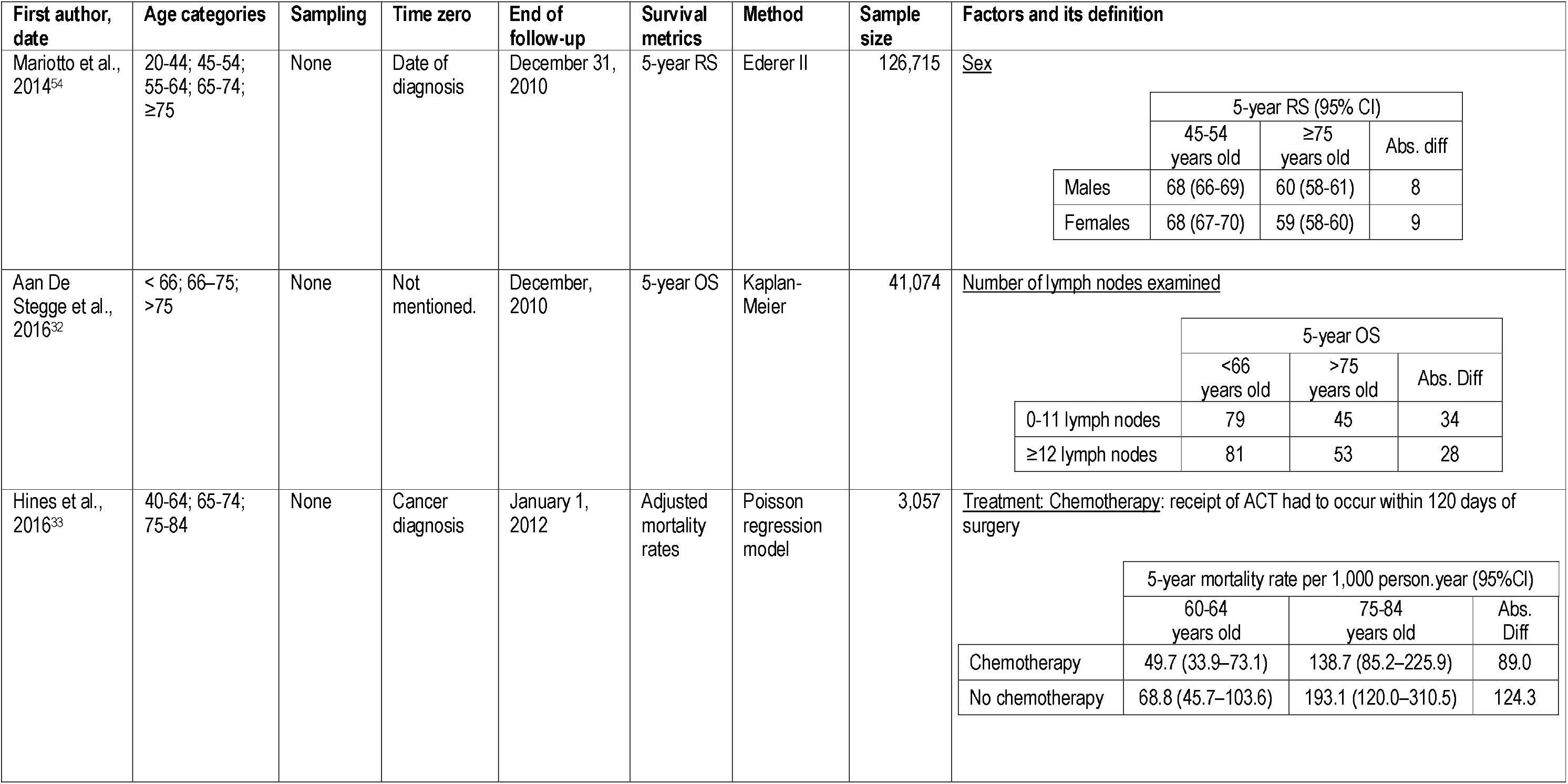
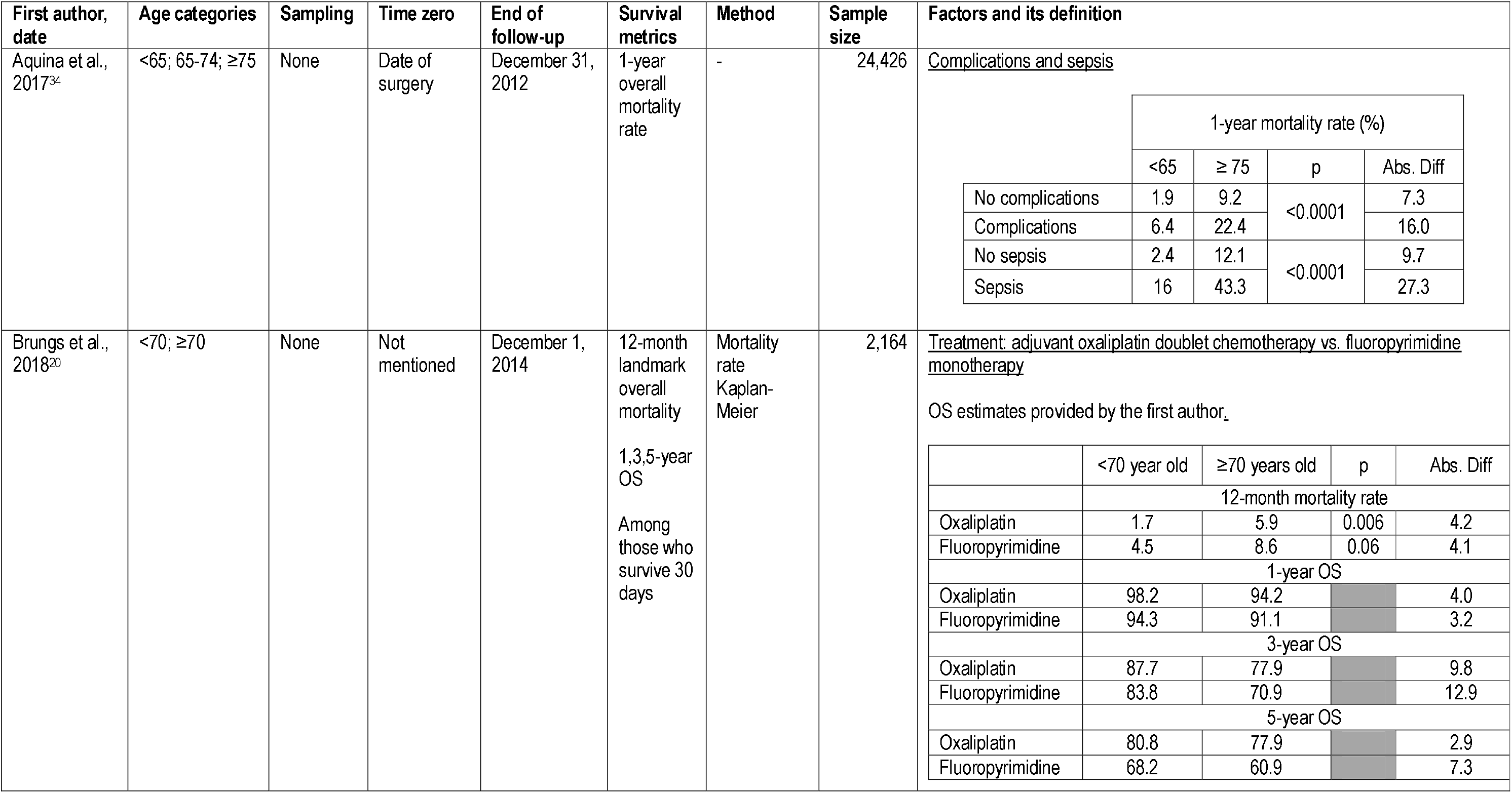
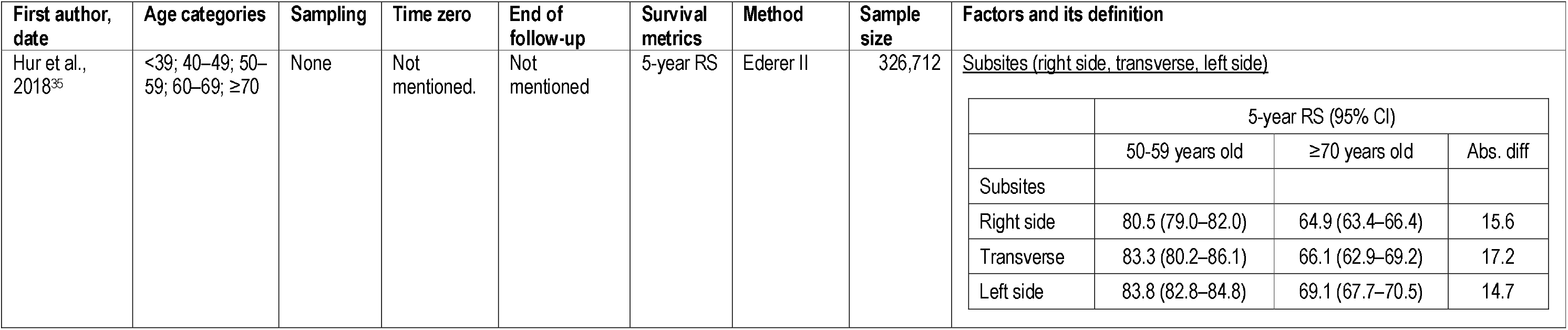
Data extraction of included studies about age disparities in colon cancer survival

**Table 5.**
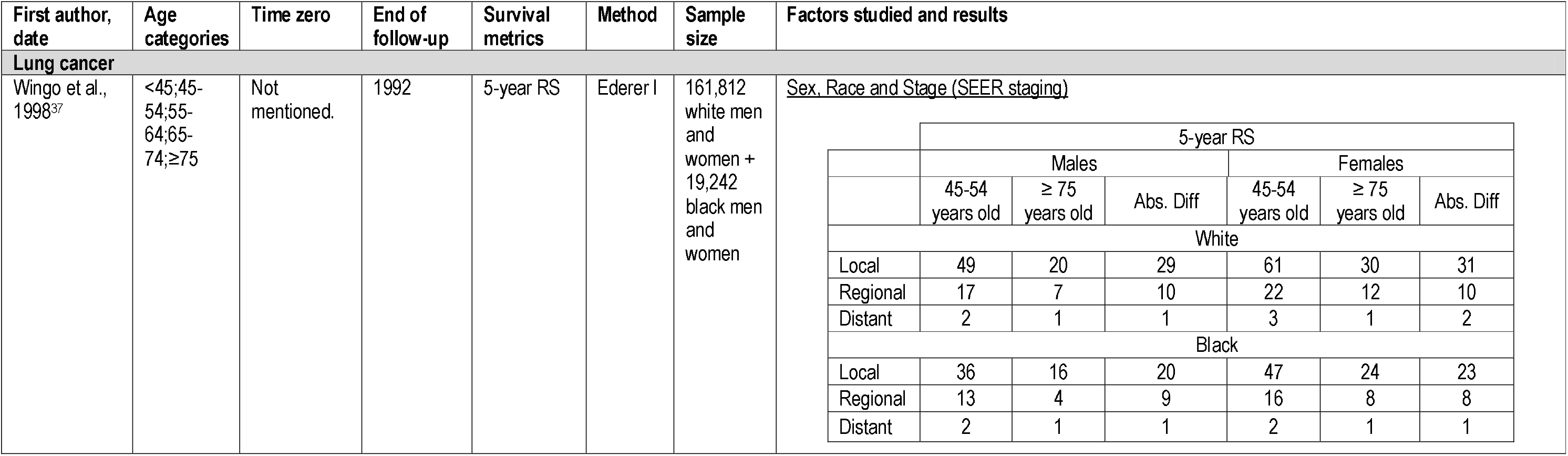
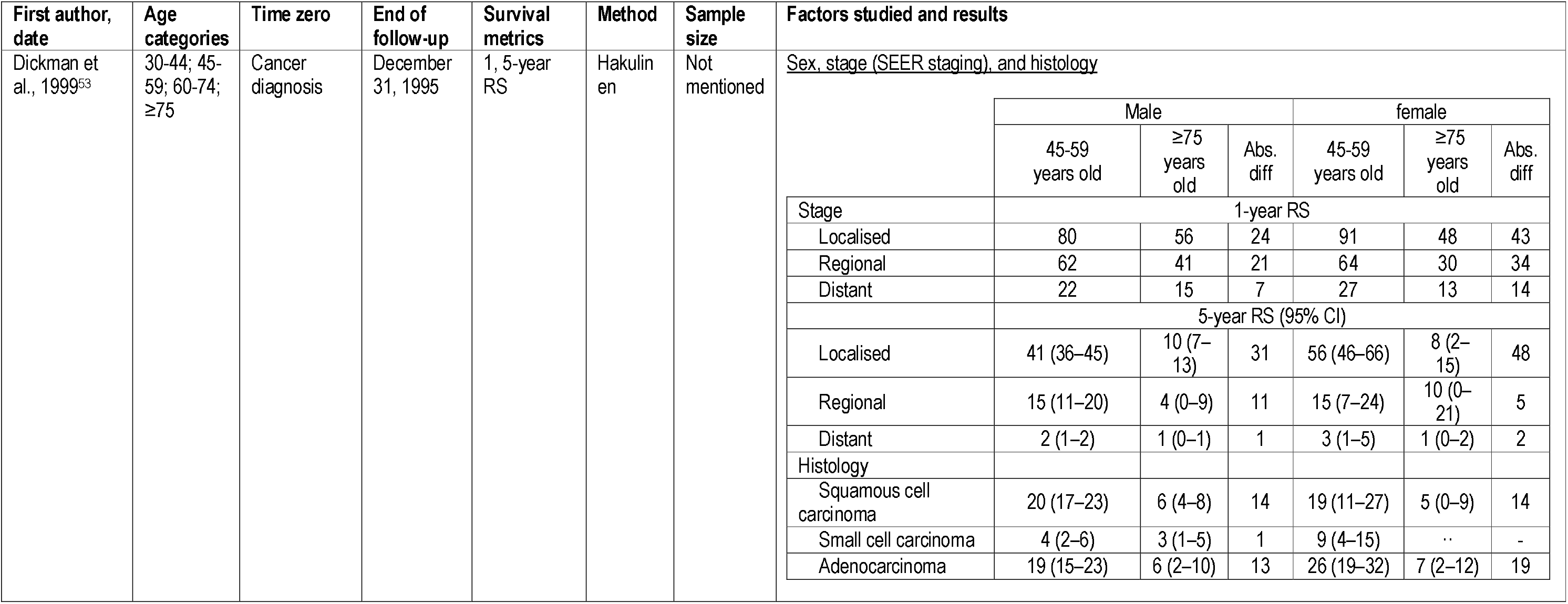
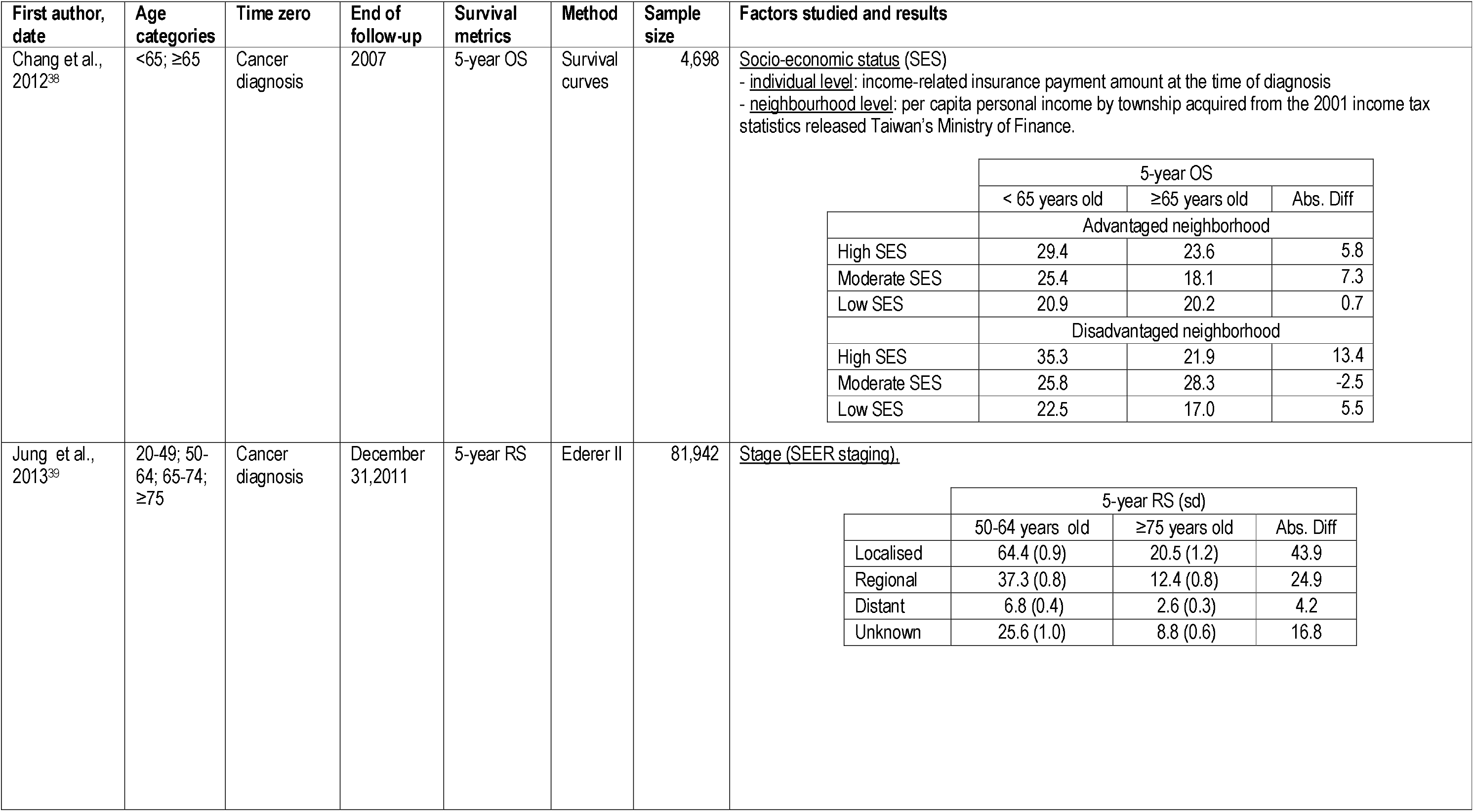
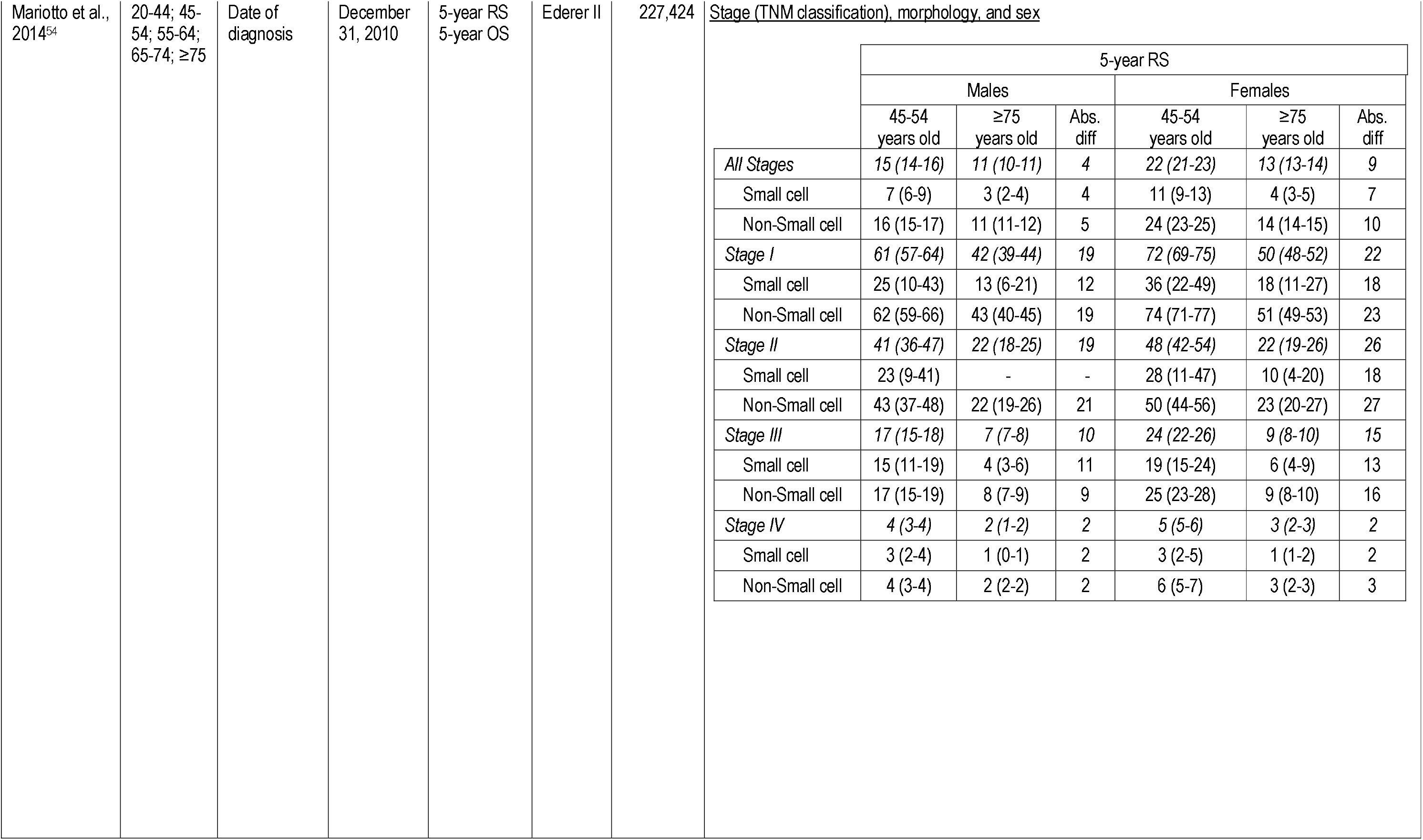
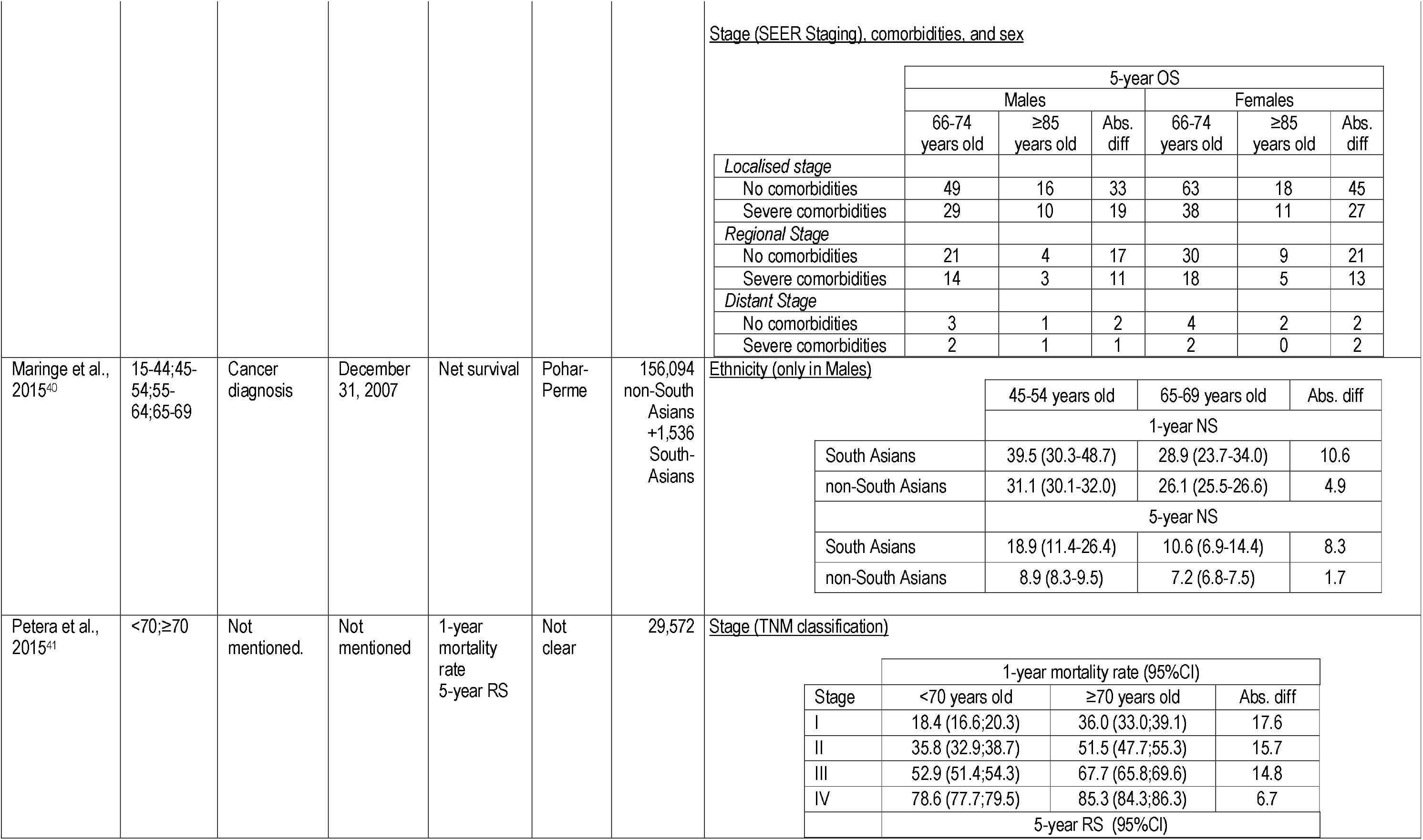
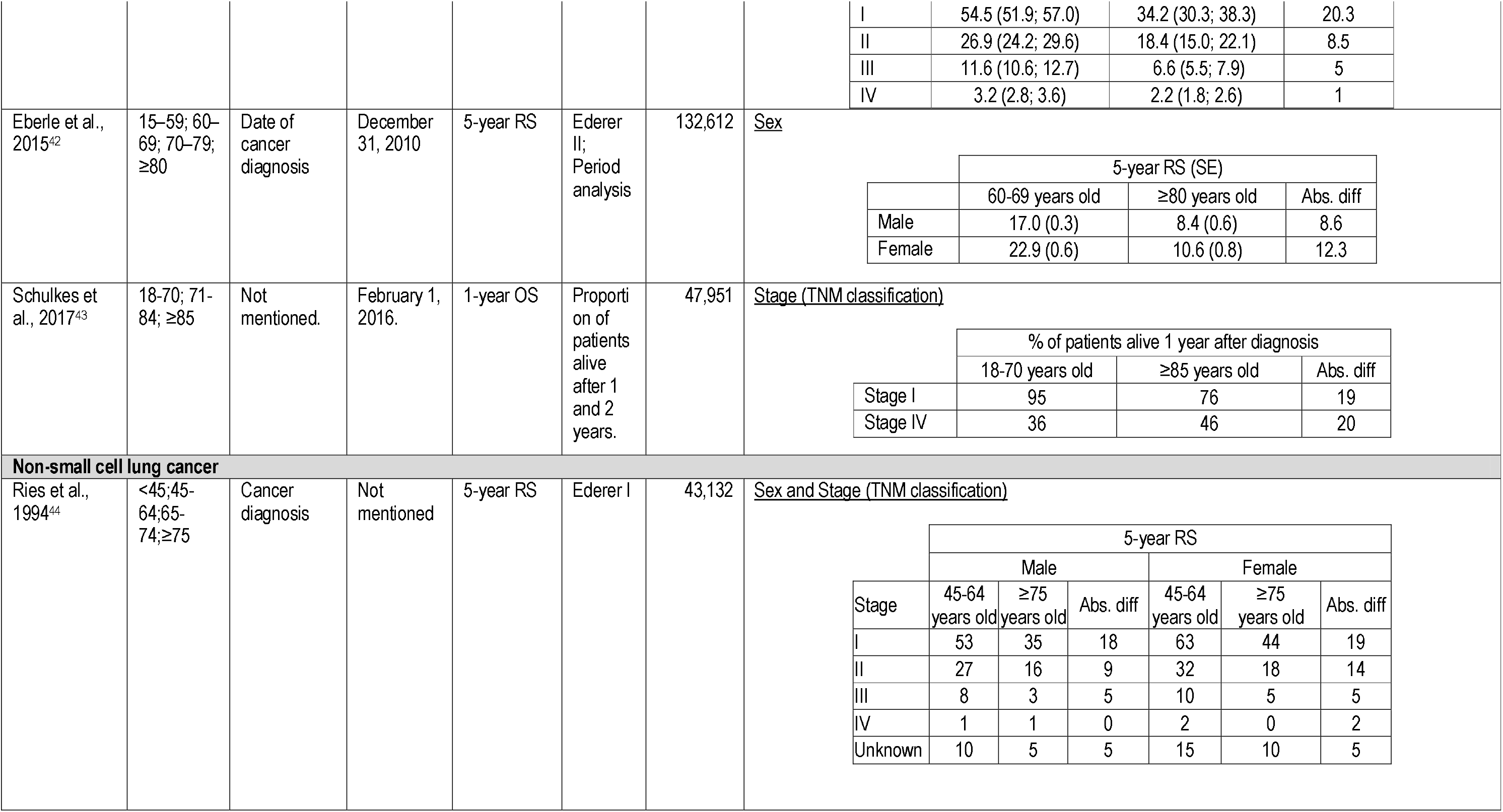
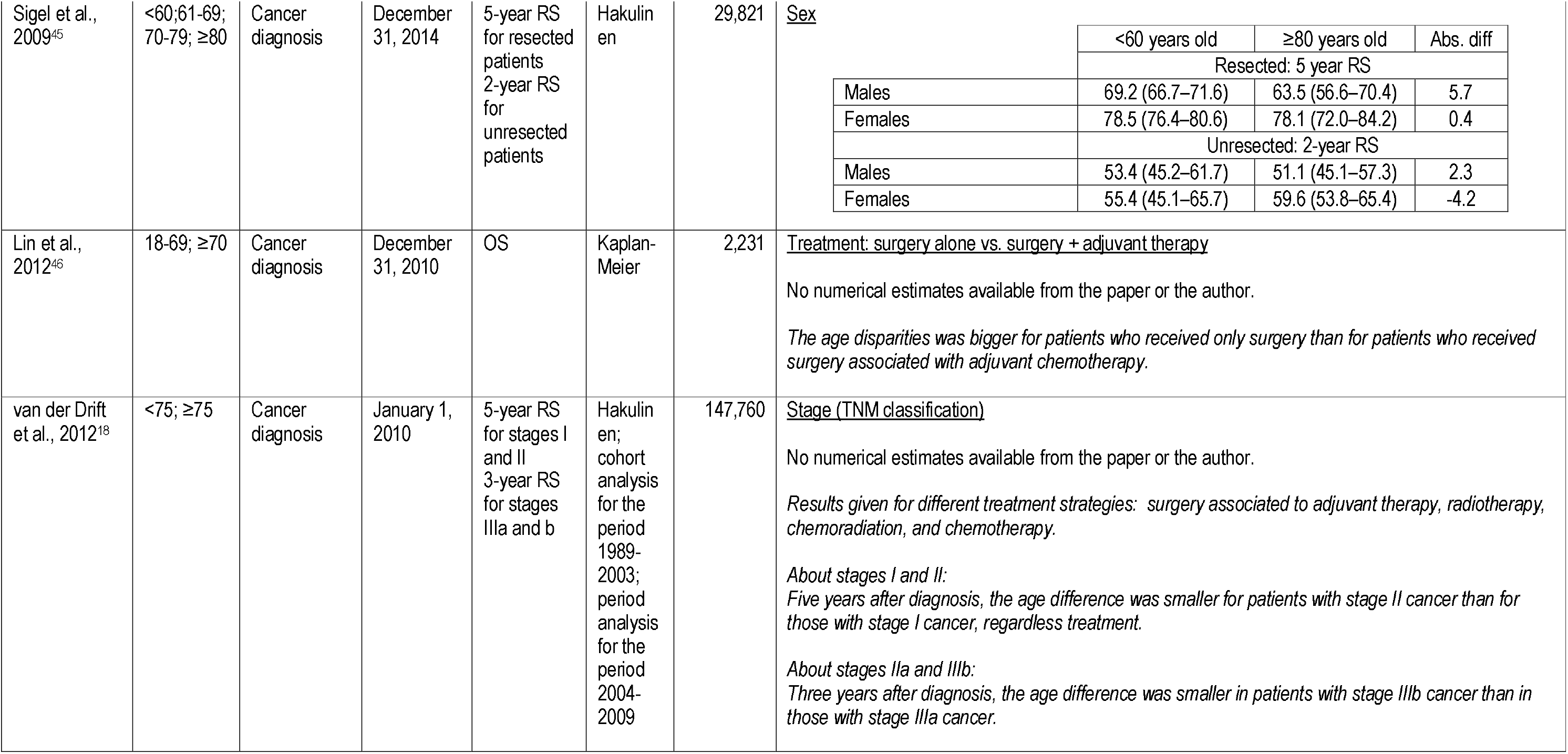
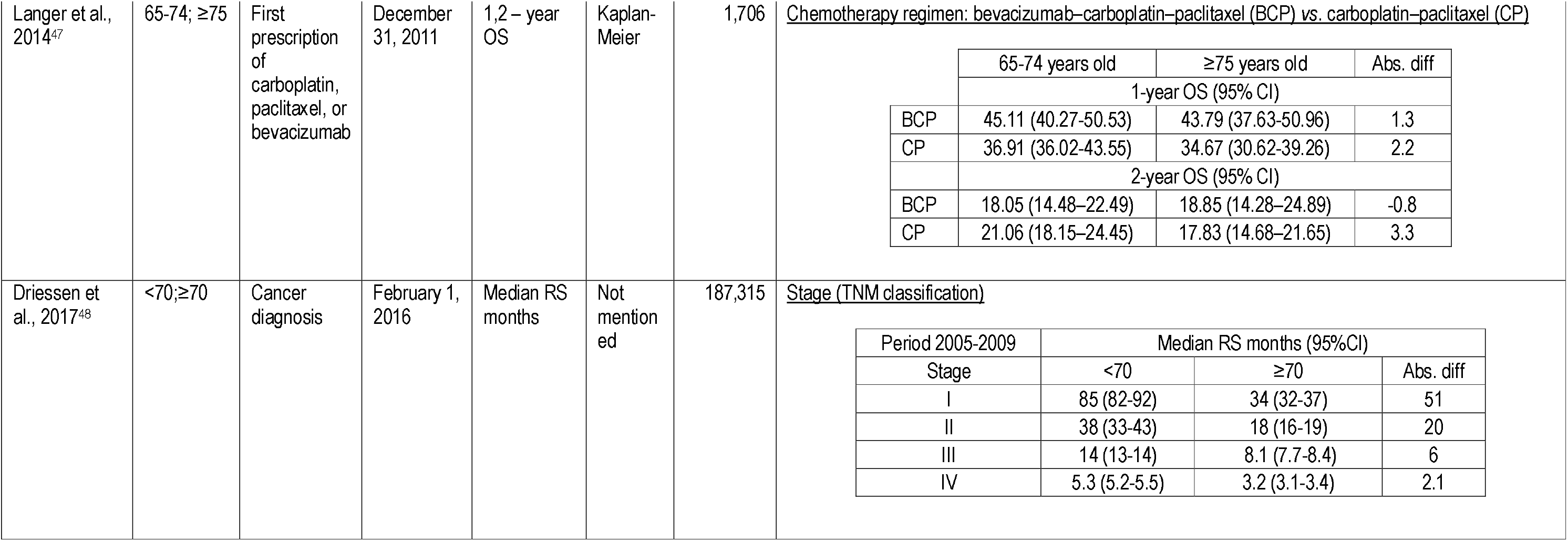
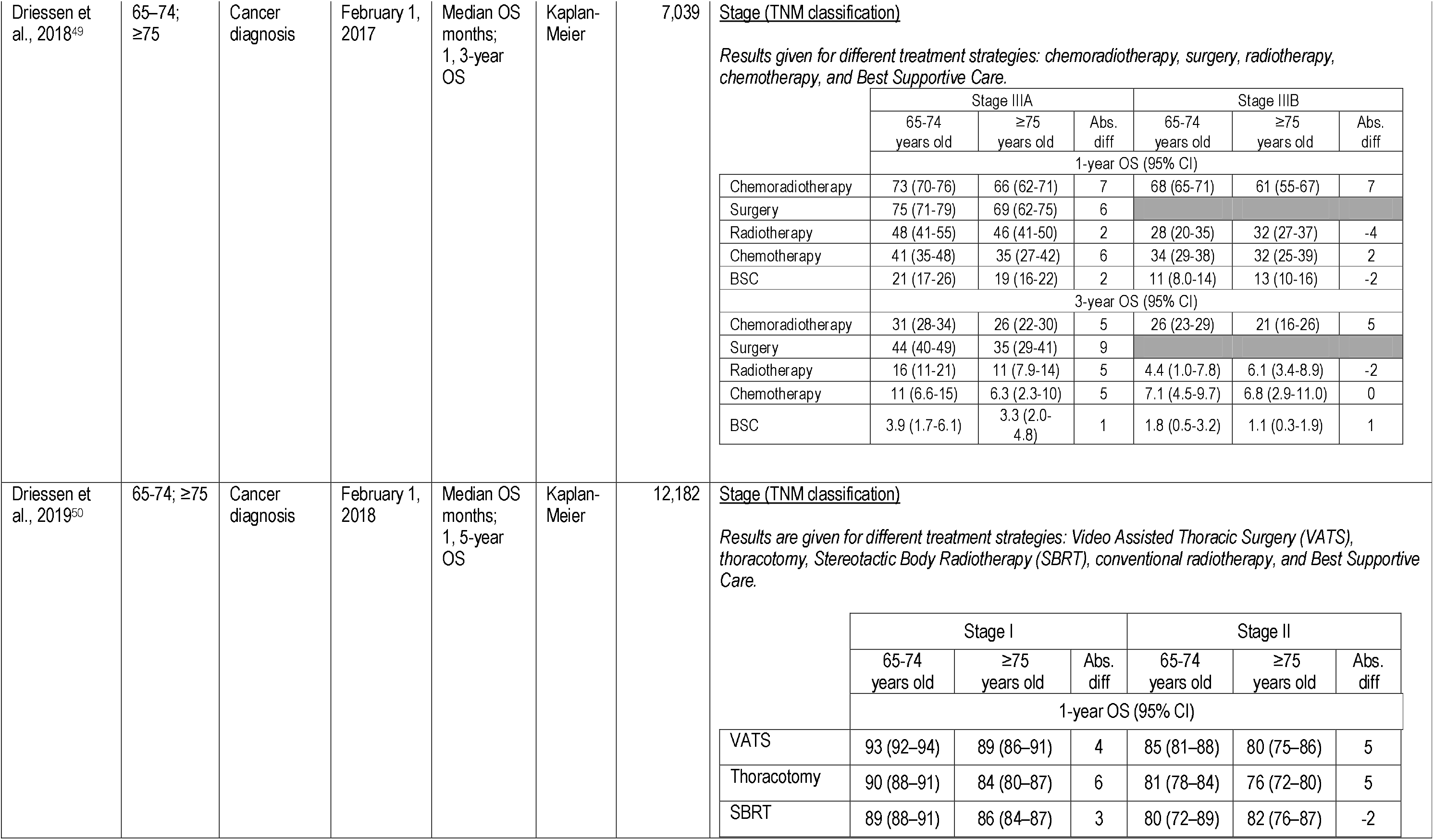
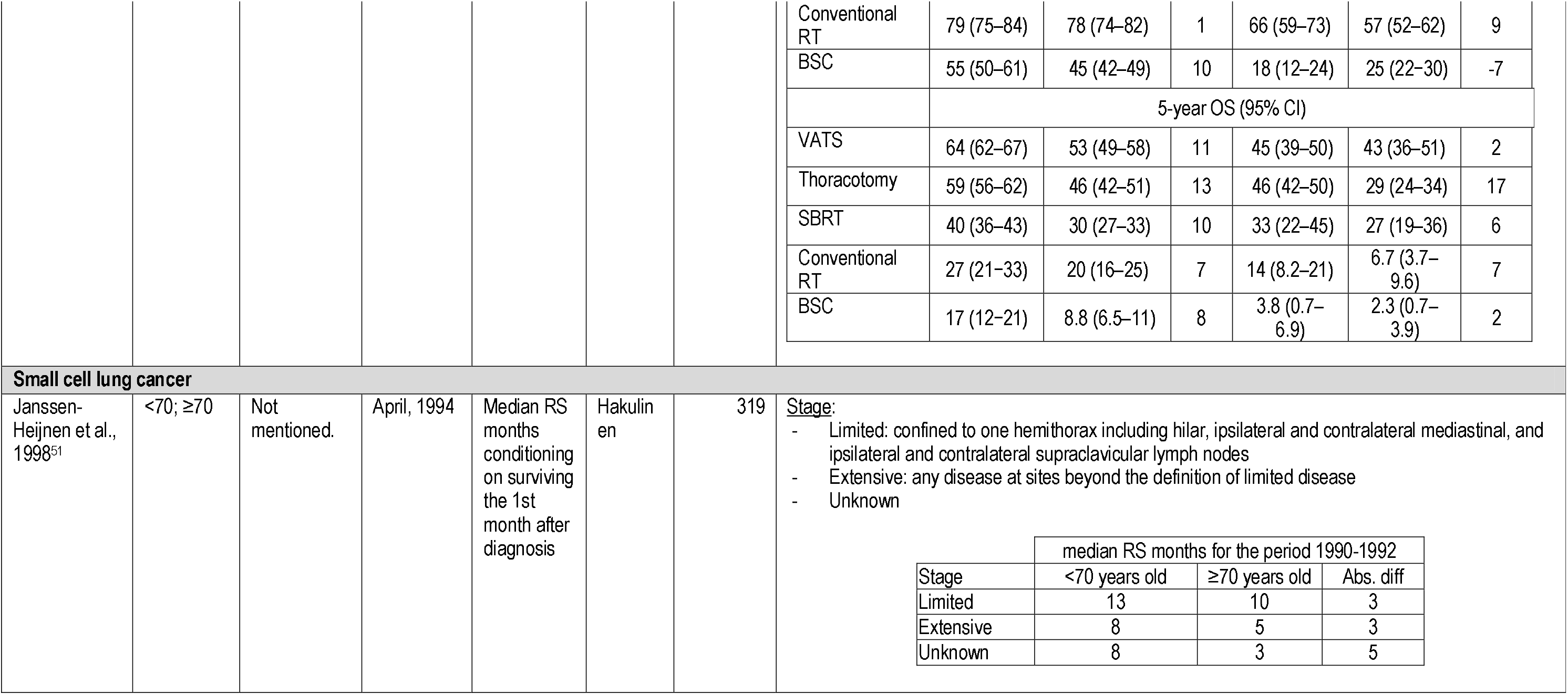
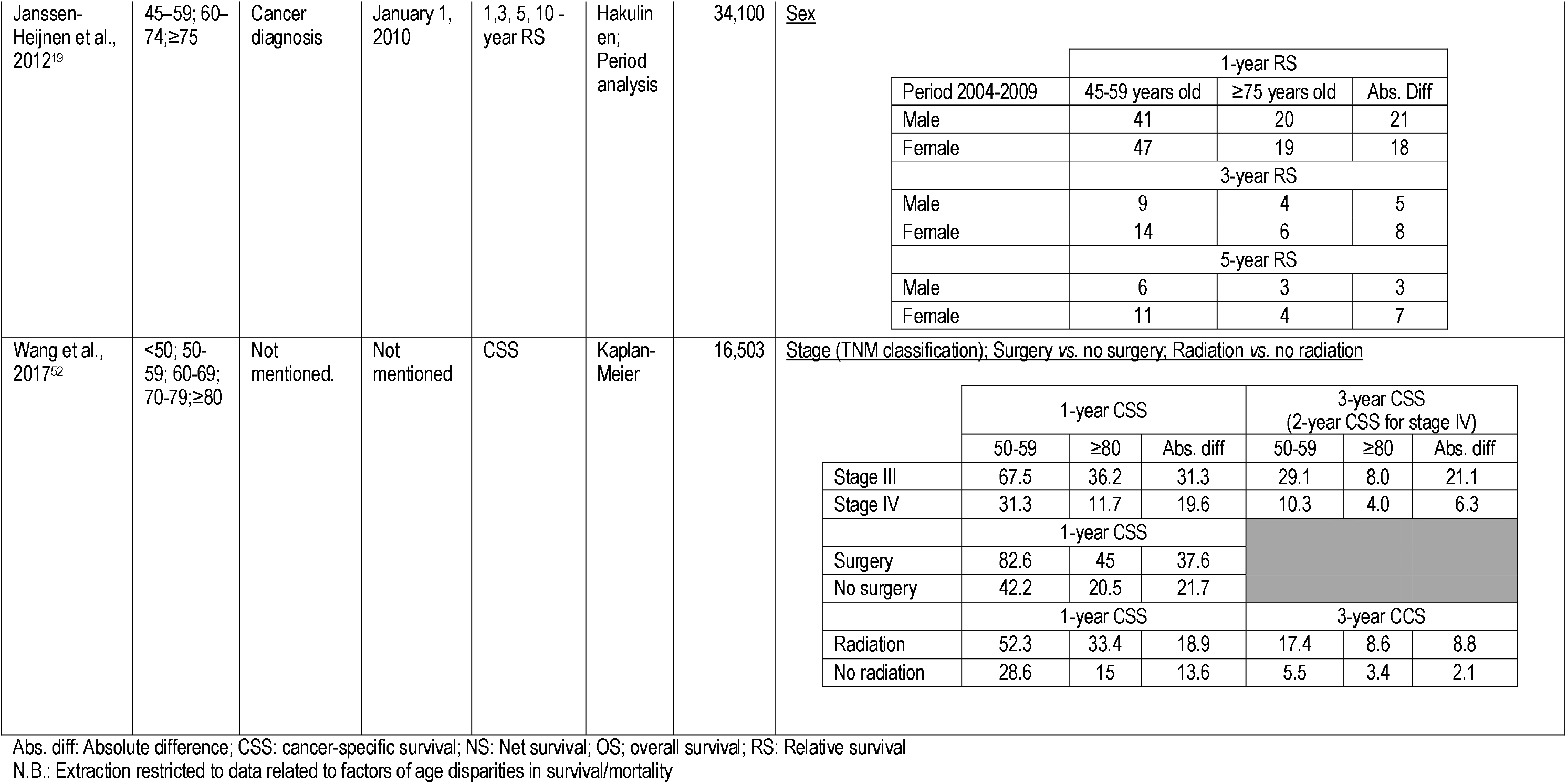
Data extraction of included studies about age disparities in colon cancer survival

Of the twenty studies that examined lung cancer, eight studies focused on non-small cell lung cancer (NSCLC)^18,44–50^, three on small-cell lung cancer (SCLC)^19,51,52^, and nine on all lung cancer cases^37–43,49,53,54^ **(Table 3)**. Six studies aimed to evaluate age disparities in surviva1^41,43,45,48–50^. Seven studies analysed data from the Netherlands^18,19,43,48–51^, six from the U.S.A^37,44,47,52,54^, and the remaining studies presented data from Finland^53^, Taiwan^38,46^, Korea^39^, the Czech Republic^41^, England^40^, and Germany^42^. While most studies included all stages at diagnosis, some studies restricted their sample to specific stage(s): stage I cancer^45^, stages l-llla^46^, stages IIIb and IV^47^, stage III^49^, and stages I or II^50^. Ten studies included patients of all ages at diagnosis, other studies included patients from the age of 15 (n = 3)^18,40,42^, 18 (n = 2)^43,46^, 20 (n = 2)^39,54^, or 65^47,49,50^. All studies used age categories that differed largely across studies in terms of number and boundaries. Twelve studies presented RS estimates^18,19,37,39,41,42,44,48,51,53,54^, seven OS estimates^38,43,46,47,49,50,54^, one study net survival^40^, and one study presented cancer-specific survival (CSS) estimates^52^.

**Table 3.**
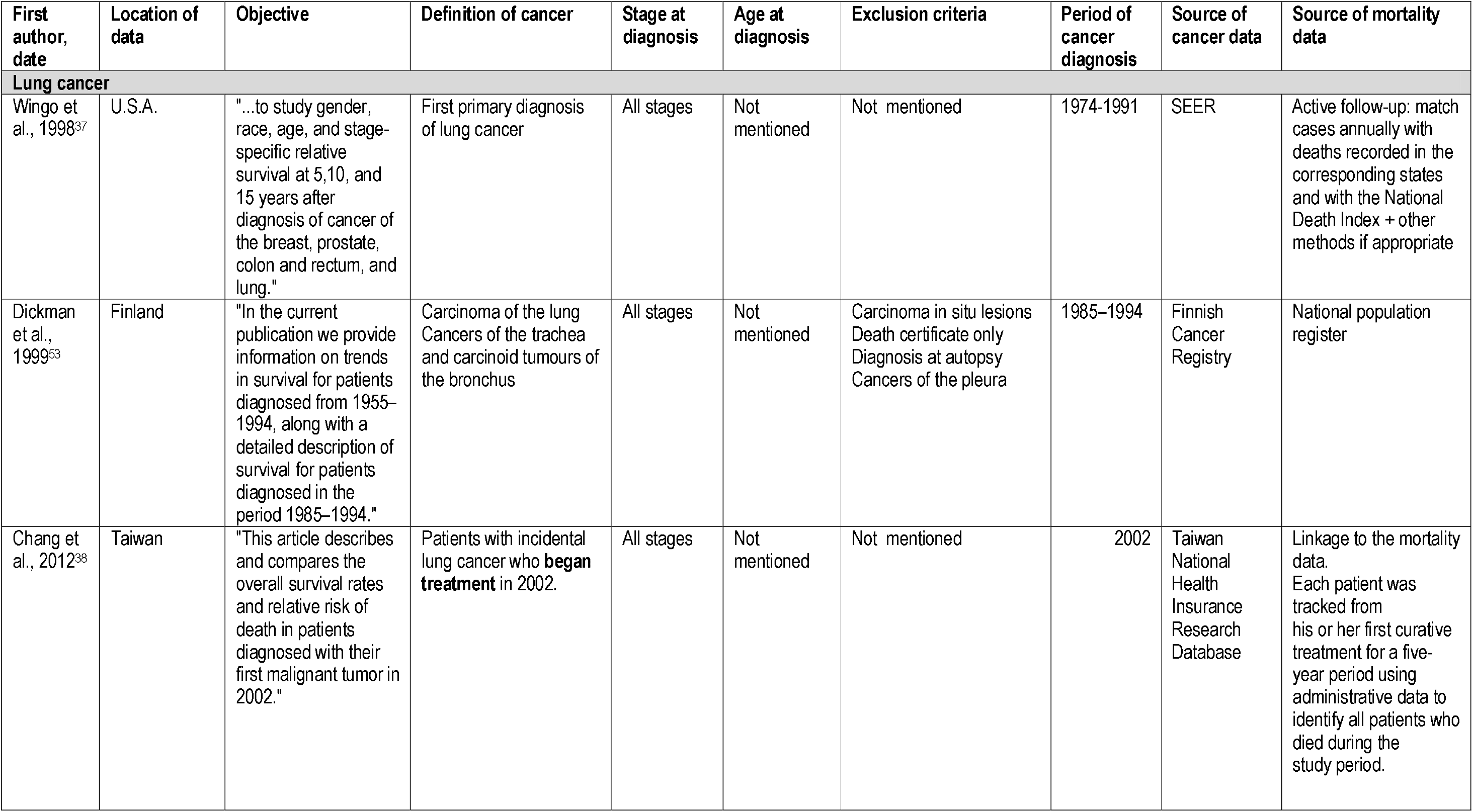
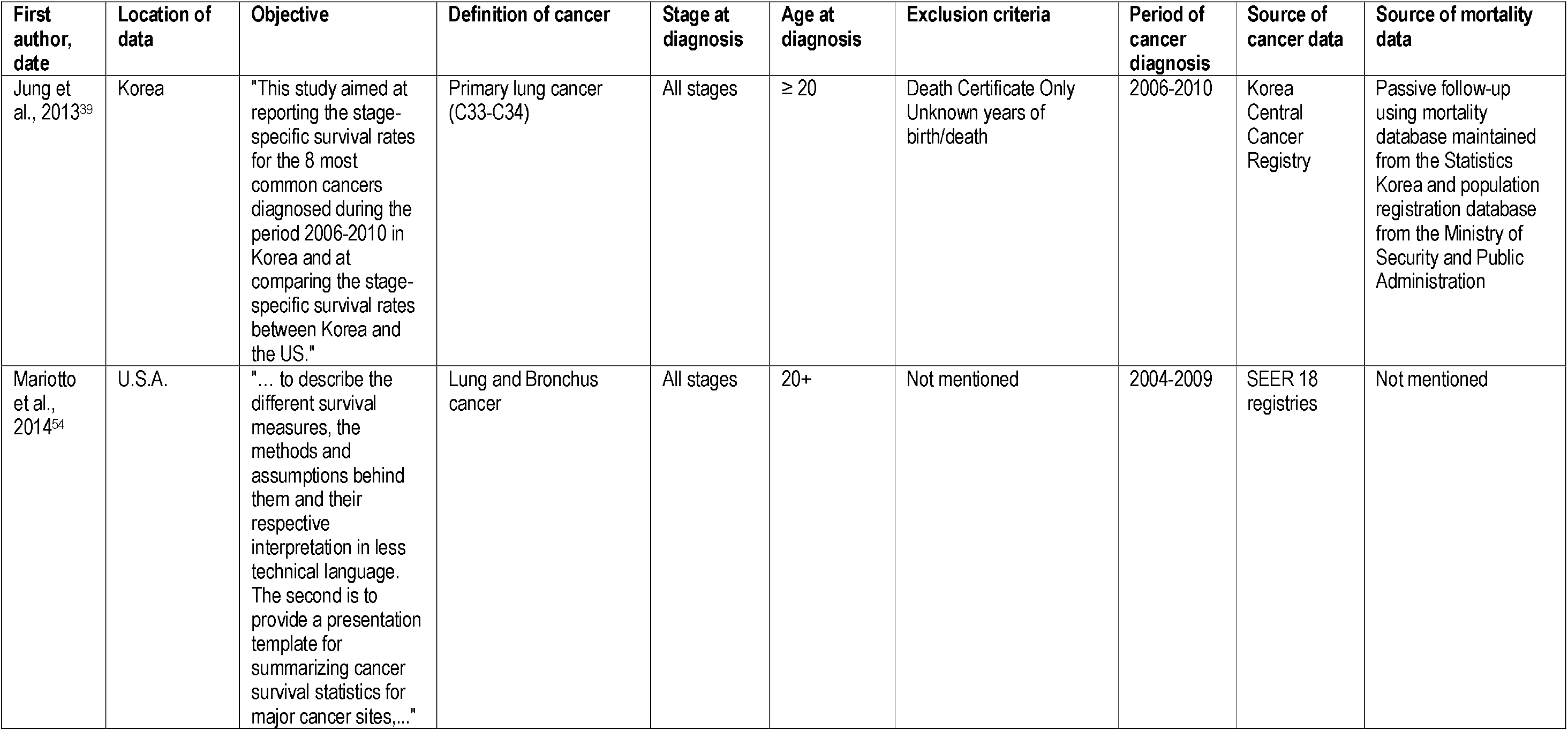
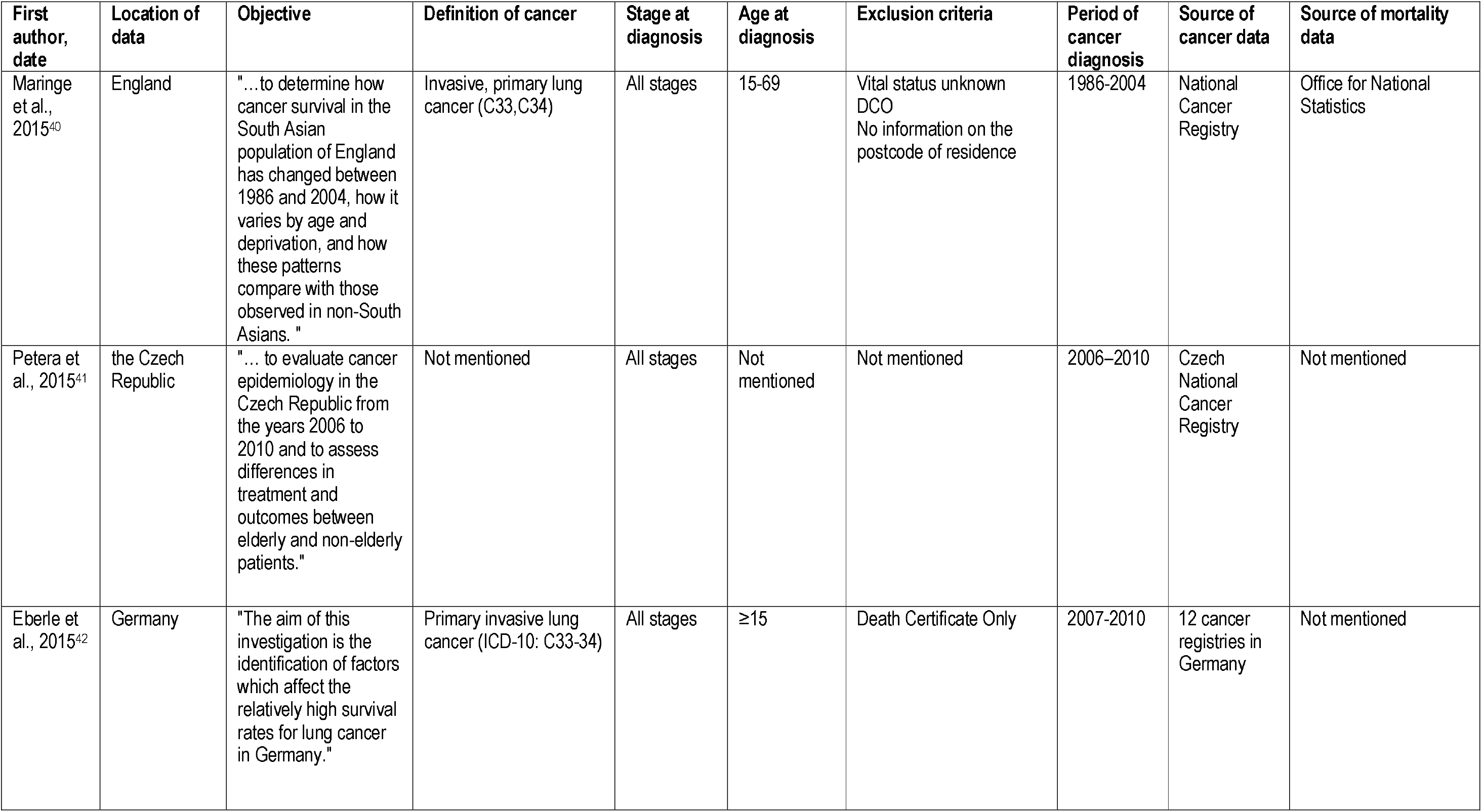
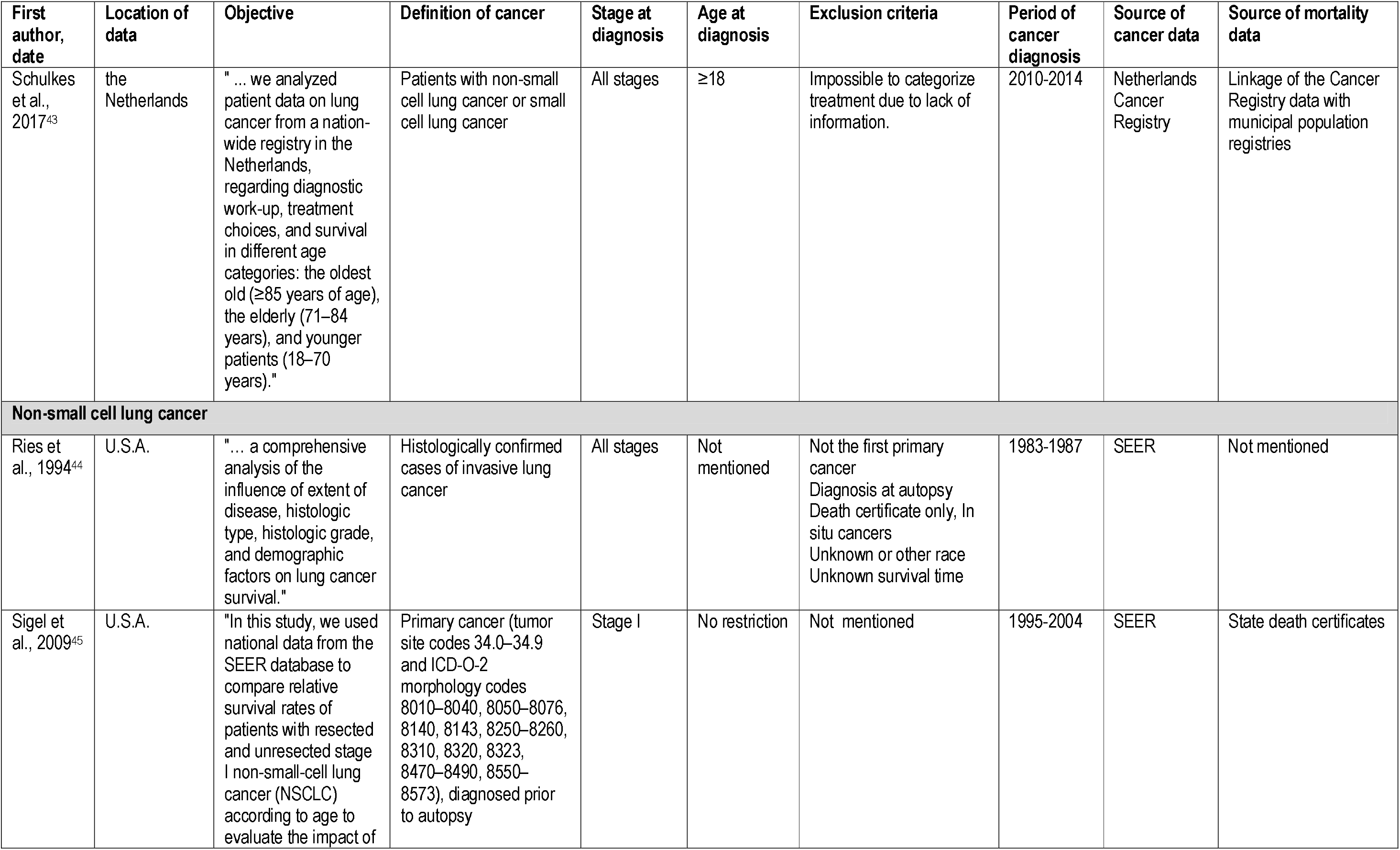
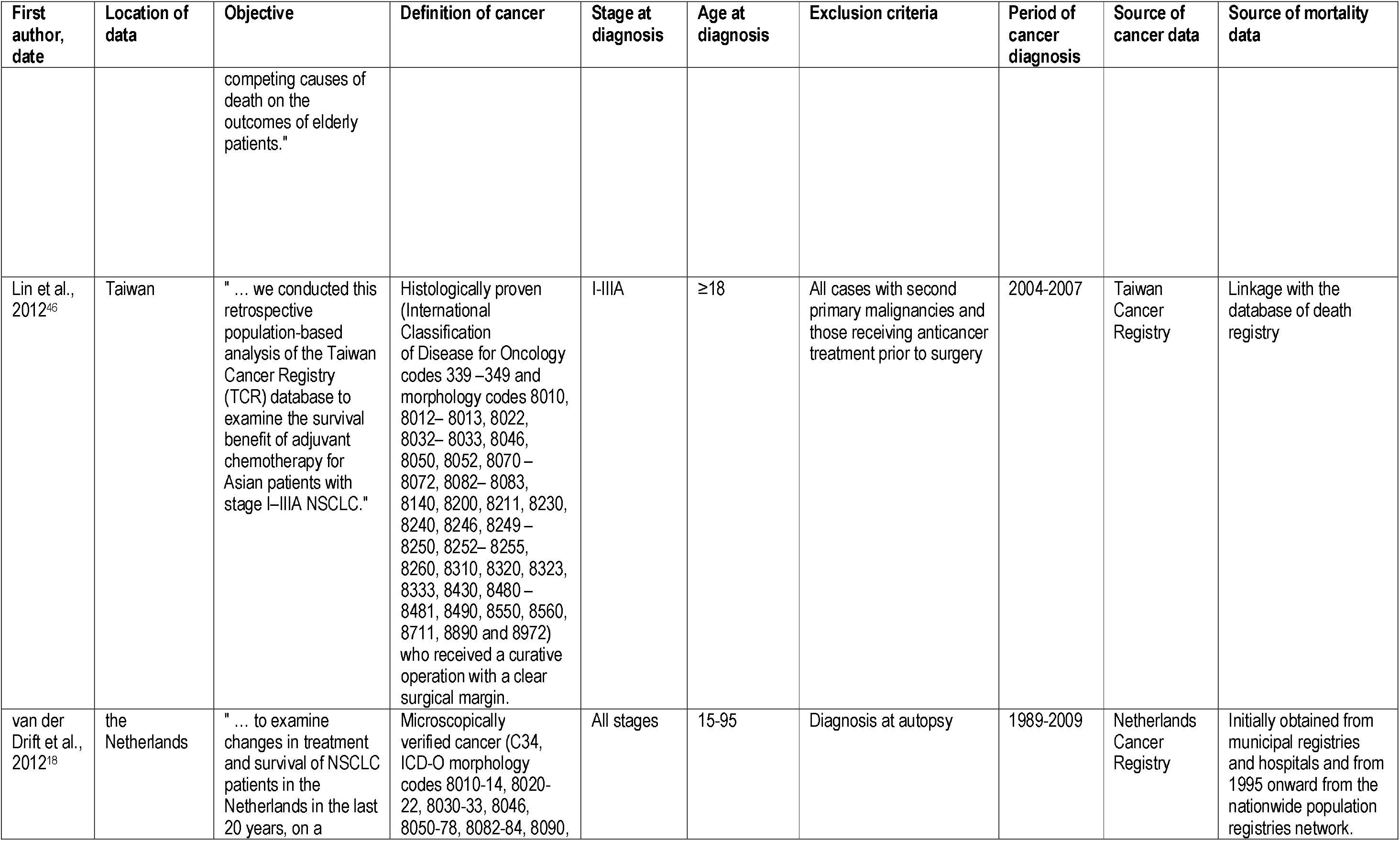
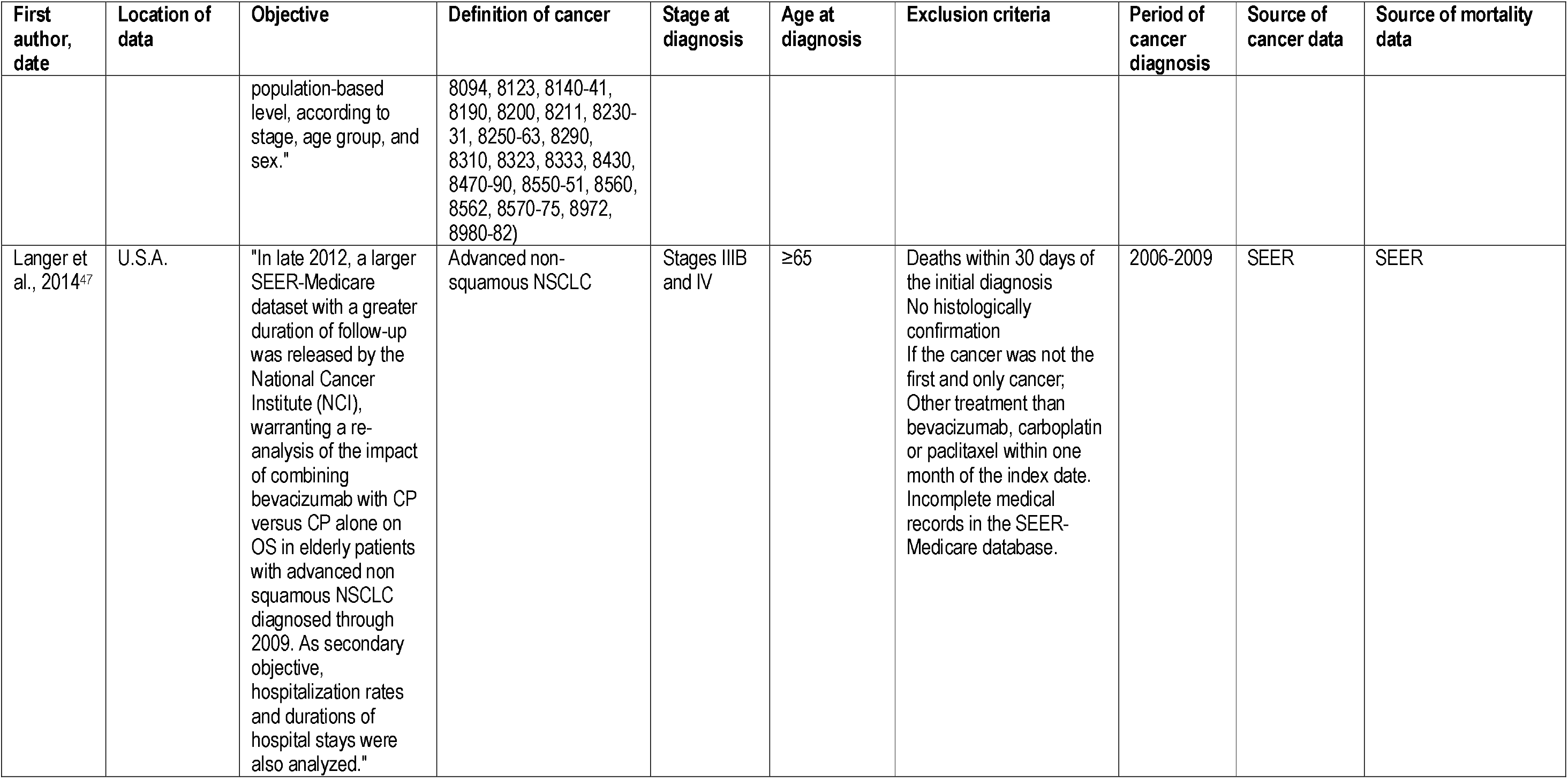
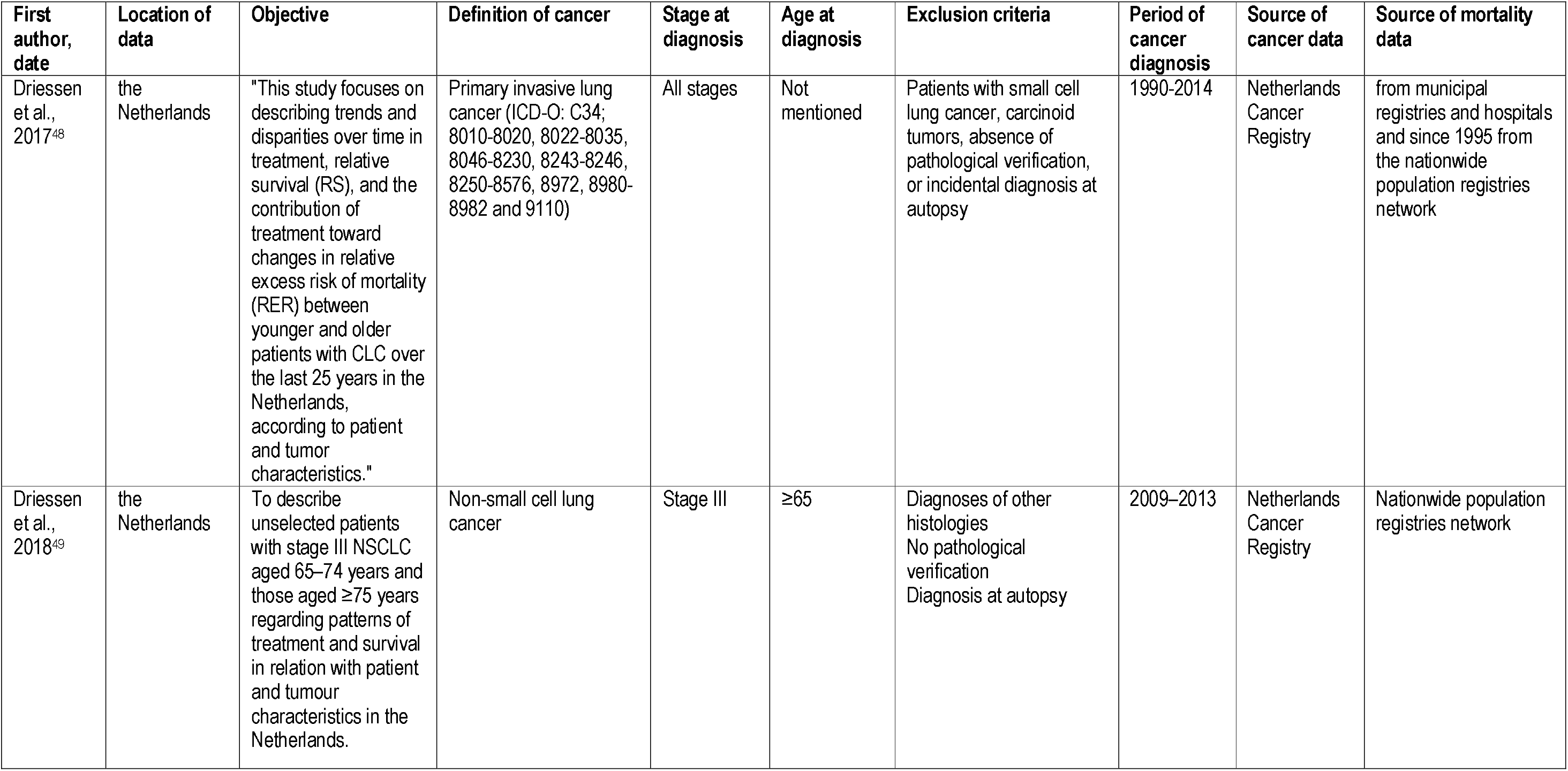
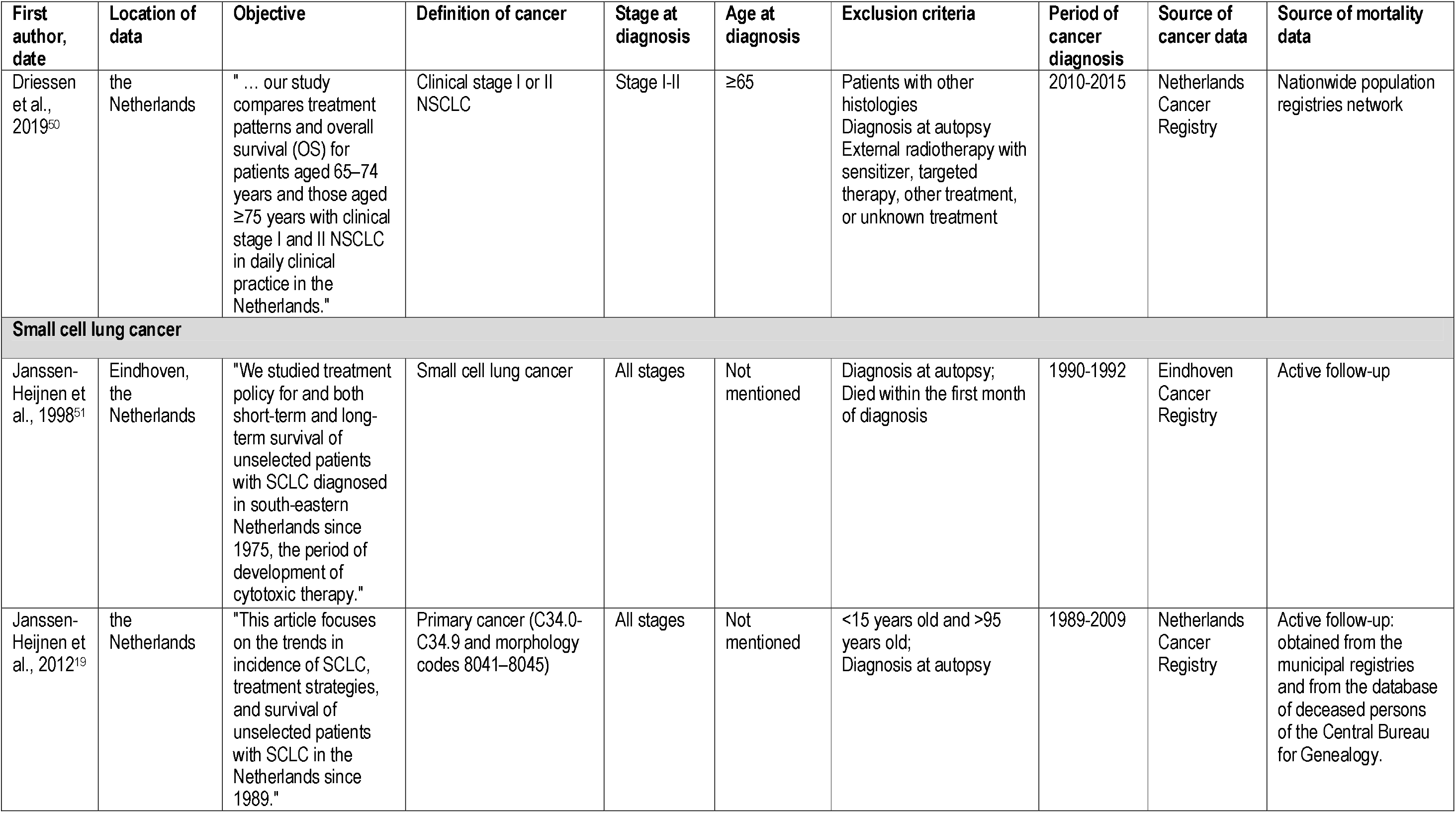
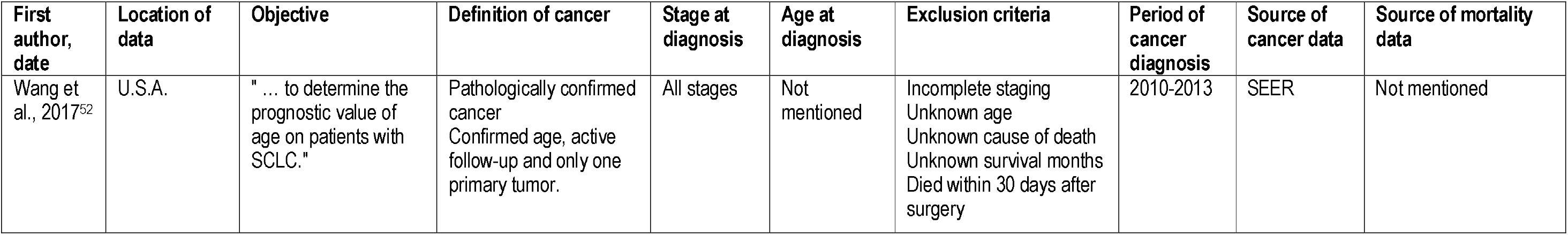
Characteristics of included studies about lung cancer

### Age pattern in colon and lung cancer survival

Patterns of age disparities in survival for colon and lung cancers based on patient-related and clinical factors are shown in **Tables 4** and **5**, respectively. All numerical values extracted from included studies are shown in **Supplementary Tables 5** and **6**. While a detailed breakdown of key findings on patterns of age disparities from our review is shown in **Supplementary Material x**, we have summarised key findings below.

Regarding colon cancer survival, higher age disparities were observed in females with regional or distant cancers^53^, and those with left colon cancer^29^, while the other studies did not^54^. Overall, age disparities were greater as cancer spread or when the cancer stage was unknown^16,26,30,53^, when lymph nodes were involved^28^, or when fewer than 12 nodes were examined^31,32^. While some studies did not show different age patterns in survival based on sub-site^35^, others reported smaller age differences for patients with cancer of the distal colon compared to the proximal colon^36,53^. Regarding treatment, the presence of bias precludes accurate interpretation of studies that presented survival data across chemotherapy regimens^26,27,33^. One study reported postoperative mortality rates in patients who underwent an elective and non-elective resection^17^. It showed higher age disparities for males, those with an American Society of Anesthesiologists (ASA) score of ≥ 3, those with a Charlson comorbidity score ≥2, those with metastatic disease, and those with hemicolectomy. Complications and sepsis after surgery^34^, as well as the presence of chronic obstructive pulmonary disease at the time of cancer diagnosis^21^, would also likely increase age disparities in colon cancer survival.

Regarding lung cancer survival, females had consistently higher age disparities in relative survival^19,42,44,45,53,54^. We observed no clear pattern for the role of socioeconomic level on age disparities^38^. Regarding the role of race/ethnicity, one study reported smaller age disparities in lung cancer survival among Black patients compared to White patients in the U.S^37^. In comparison, South Asians had higher age disparities than non-South Asians in the U.K^40^. Age disparities tended to decrease as the cancer spread^37,39,41,44,48,50–54^ and were higher in patients with NSCLC than in those with SCLC^53,54^. One study suggested that that age disparities were smaller in patients with severe comorbidities than in those without comorbidity^54^. Again, most studies presenting survival data by treatment group were at high risk of bias^18,46,49,50,52^. The only interpretable study showed that age disparities in overall survival did not differ based on the chemotherapy regimen^47^.

## Discussion

This review is the first to bring together the literature on those factors which influence age disparities in cancer survival, using colon and lung cancer as exemplars. While age at diagnosis is an important prognostic factor in cancer survival, very few studies, often of sub-optimal quality, have specifically focused on the relationship between age and cancer survival, and none have sought to identify patterns of age disparities in colon or lung cancer survival *per se*. However, our review showed that i) the magnitude of disparities in survival between younger and older patients differed greatly and inconsistently based on patient and clinical characteristics; ii) the stage at diagnosis was the sole clinical characteristic that consistently influenced age disparities in survival, but oppositely in colon cancer and lung cancer; and iii) age disparities in lung cancer survival were typically greater in female than in males.

While in most studies older patients had poorer survival than middle-aged patients, this was not always the case. For instance, two studies reported no age disparity in cancer survival in patients with cancer of the right colon^53^, and other papers showed minimal age disparities in patients with advanced lung cancer^29^, or small-cell lung carcinoma^53^. On the other hand, age disparities were substantial in patients with distant colon cancer^30,53^ or those with localised lung cancer^37,41,44,48,53,54^, particularly for patients without comorbidities^54^.

The influence of stage at diagnosis on age disparities differed depending on the cancer. Age disparities in colon cancer survival tended to increase with increasing stage of disease^28,30^, while the opposite was observed for lung cancer^37,41,44,48,53,54^. Surgery is the main treatment strategy for patients with colon cancer diagnosed with localised and regional stage disease, while chemotherapy is recommended for metastatic colon cancer^57^. It has been shown that older patients are less likely to receive chemotherapy than younger patients^58–60^, and less intensive therapies are usually recommended for unfit older patients^61^. In lung cancer, older patients with early stage disease, especially those older than 75, are less likely to undergo surgery compared to younger patients^62^. The high lethality of the disease, especially at a more advanced stage, may explain the small difference in survival disparities observed between middle-aged and older patients with metastatic lung cancer.

Comorbidity, the prevalence of which drastically increases with age, is an important prognostic factor in patients with cancer, because it may complicate cancer management^63^. However, our review identified only two studies reporting data for comorbidity, preventing us from making any conclusions regarding comorbidity and its impact on age disparities in cancer survival. One study suggested that the presence of COPD at diagnosis may increase age disparities in survival seen in patients with colon cancer^21^. The sole study that presented lung cancer survival data by age group and comorbidity level suggested that patients without comorbidities showed greater age disparities in survival than those with severe comorbidities^54^. Comorbidity alone is not enough to assess vulnerabilities in older patients with cancer, and comprehensive geriatric assessments (CGA) may be useful in capturing a more nuanced view of health, fitness and physiological aging^64^. Although less valuable than information derived from CGA, it is now possible in many countries to link cancer survival data to comorbidity information through linkage with administrative hospitalisation data or pharmacy data^65,66^, and thus further studies should describe the role of comorbidities on age disparities in survival in patients with colon or lung cancer.

Unfortunately, we are unable to draw any conclusions regarding the role of treatment on age disparities in colon and lung cancer survival. Indeed, most studies presenting survival data by treatment group were at high risk of immortal time bias^27,33,49,50,52^. Immortal time bias occurs when survival comparisons are made between groups of patients based on a factor (e.g. treatment) that is defined after the start of the follow-up (e.g. cancer diagnosis date). Patients in the treated group survived long enough to be treated, while others in the untreated group may have died before having that chance. As a consequence, the treatment may be erroneously considered as effective because patients in the treated group have, on average, a better survival than those in the untreated group. In reality, the apparent better survival in the treated group may be the result of the selection of the fittest patients *[i.e*. those who had the better chance to survive). For instance, this bias may be at play in the 2010 study of van Steenbergen et al. and would explain the higher survival amongst the oldest age group in the ‘no chemotherapy’ group^27^, or in the study of Sigel et al. that reported higher 2-year RS in female patients older than 80 years compared to those younger than 60^45^. With a few exceptions^34,40,47,53,54^, the overall quality of studies included in this review was poor. Further high quality studies are required if we are to better identify the role of treatment as a possible driver of age disparities in cancer survival.

Only a few studies provide information about patient characteristics. The sole patient characteristic examined in the colon cancer studies was sex, and the results were inconsistent^29,53,54^. However, the included lung cancer studies suggested that the difference in 5-year survival between younger and older patients is wider in females than in males^19,37,42,53^ but this was not necessarily the case for 1 and 3-year survival^19^. In the study by Dickman et al., females aged 45-59 years old had better 1-year RS than males of the same age; however, females aged 75 years or older had lower 1-year RS than their male counterparts^53^. Even if some evidence suggests a positive effect of sex hormones on survival from NSCLC in females^67^, the implication of sex hormones is still not Clear^68^. However, because of the observational nature of the studies included, survival bias may also be an explanation for the difference observed across sexes. In terms of race/ethnicity, age disparities in lung cancer survival seem to be influenced by race/ethnicity in the U.S and the UK, but results are inconsistent^37,40^, probably because of differences between health-care systems, or possible survival bias. Finally, the role of socioeconomic level in age disparities in lung cancer survival is not Clear^38^. While sex, ethnicity/race and socioeconomic level are known to influence cancer survival^69–71^, their role in age disparities in cancer survival remain unclear and should be further explored.

Many other characteristics may be important in explaining lower survival amongst older patients. When using observational data, data related to demographics and cancer are the easiest to study. However, other factors such as those related to the health-care system (e.g. physical and financial access to cancer facilities) are likely to be more difficult to measure, and therefore were less likely to be included in this review.

Older adults have a higher risk of dying from causes other than cancer than younger adults. While of interest for patients and clinicians^72^, OS measures are of limited value when studying disparities in survival between younger and older patients, mainly because they do not make a distinction between the causes of death, and because of the higher risk of background mortality in older patients. Identifying the underlying cause of death may be challenging in older adults who may present with co-existing serious disease, making cancer-specific survival difficult to estimate. When studying the age disparities in survival, it is therefore crucial to take into account this difference in background mortality. Accordingly, relative survival (*i.e*. the ratio of the observed survival among patients with cancer over the [expected] survival among the general population obtained from national life tables) or net survival (*i.e*. the probability of being alive after a defined period of time in the hypothetical world where one can die only from cancer) are suited to this purpose. However, lifetables used to estimate the expected survival should be adequately stratified by likely important factors(e.g. comorbidity, smoking status)^73^.

Our systematic review has limitations. We could not conduct any quantitative analysis (such as meta-analysis) because of the vast heterogeneity of studies included, which prevented us from quantifying the relationship between increasing age and cancer survival. This is largely a reflection of the quality of the studies included in this review. We did, however, attempt to synthesise the available evidence into the key findings, as discussed above.

With the rapidly increasing number of older patients with cancer^14^, there is a dire need for a better understanding of the drivers of the disparities in colon and lung cancer survival between older and younger patients, ultimately enhancing the probability of patients surviving their cancer regardless of their age. While it is not realistic to believe that survival amongst older adults can equal that of middle-aged adults, there is more that can be done to minimise age disparities in colon and lung cancer survival – but the current quality of evidence prevents a full understanding of the key drivers of these disparities.

## Conclusion

In this systematic review, we have investigated age disparities in cancer survival using colon and lung cancer – two differing cancer contexts in terms of the likely impact of age on survival – as exemplars. The present review highlights both the lack of knowledge about age disparities in colon and lung cancer survival, and the absence of geriatric variables (e.g. cognition, functional status, social support, nutritional status, etc.) investigated within current population-based research. With the growth of the use of administrative health data in several (high income) countries and an increased emphasis being placed on data quality, we can expect a more accurate description of age disparities in colon and lung cancer survival in the near future and a subsequent improved understanding of what drives them.

## Data Availability

The study is a systematic review of literature. No data available.

## Author contributions

SP designed the study, wrote the protocol, screened all titles, abstracts and full-texts, and drafted the manuscript. HG screened 10% of abstracts and full-texts, extracted data from included papers, and critically reviewed the manuscript.

VS reviewed the protocol, screened 10% of titles and critically reviewed the manuscript.

JG critically reviewed the protocol and the manuscript.

MJH helped with the interpretation and critically reviewed the manuscript.

RC supervised the study, helped with the interpretation and critically reviewed the manuscript.

DS helped with the interpretation and critically reviewed the manuscript.

### Data statement

Not applicable.

## Declaration of competing interest

None

## Funding

Sophie Pilleron has received funding from the European Union’s Horizon 2020 research and innovation programme under the Marie Skłodowska-Curie grant agreement No 842817.

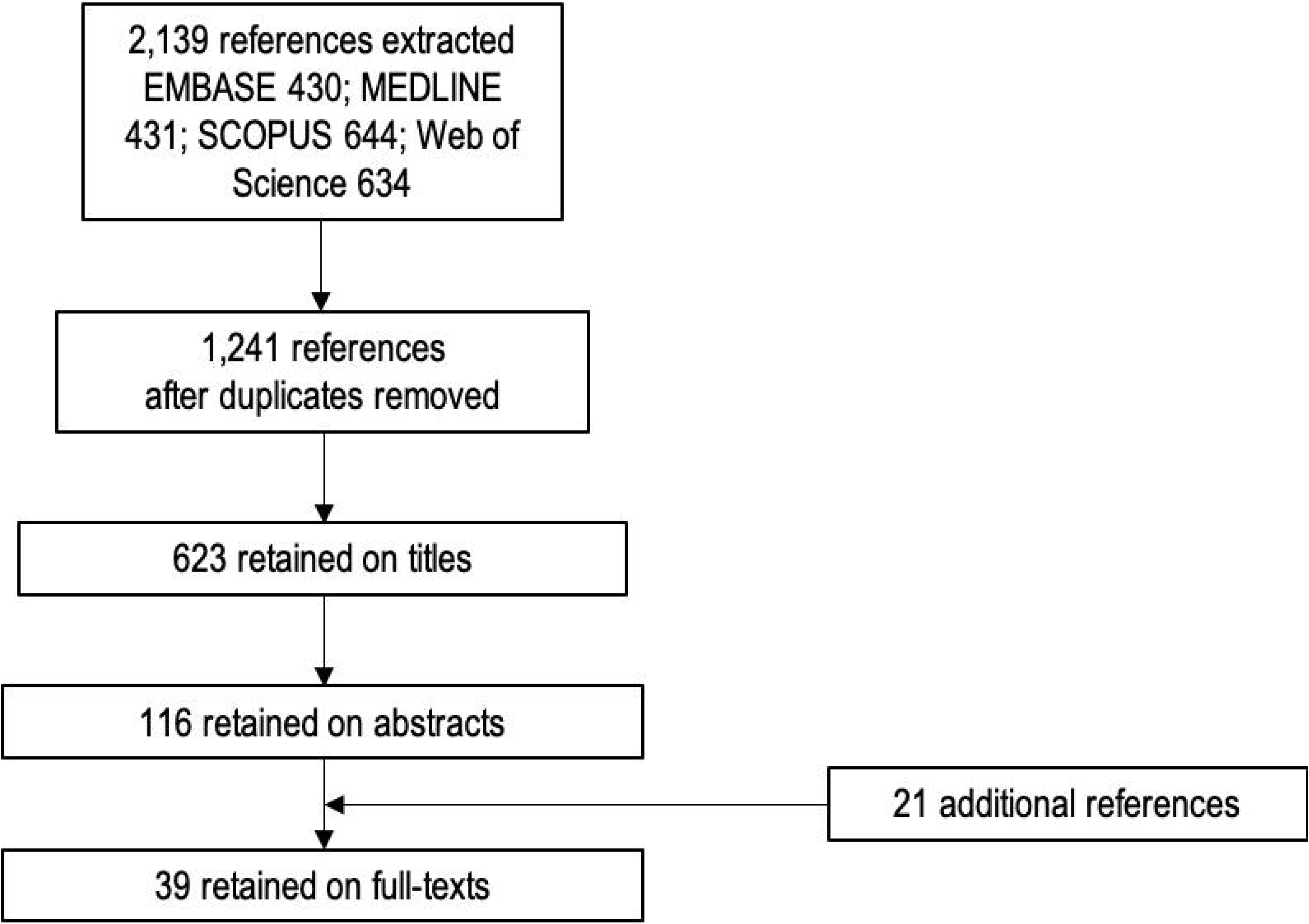

## Notes

### Competing Interest Statement

The authors have declared no competing interest.

### Author Declarations

Systematic review - No need of ethical approval

